# Mortality surrogates in combined pulmonary fibrosis and emphysema

**DOI:** 10.1101/2023.05.05.23289330

**Authors:** An Zhao, Eyjolfur Gudmundsson, Nesrin Mogulkoc, Coline van Moorsel, Tamera J. Corte, Chiara Romei, Robert Chapman, Tim J.M. Wallis, Emma Denneny, Tinne Goos, Recep Savas, Asia Ahmed, Christopher J. Brereton, Hendrik W. van Es, Helen Jo, Annalisa De Liperi, Mark Duncan, Katarina Pontoppidan, Laurens J. De Sadeleer, Frouke van Beek, Joseph Barnett, Gary Cross, Alex Procter, Marcel Veltkamp, Peter Hopkins, Yuben Moodley, Alessandro Taliani, Magali Taylor, Stijn Verleden, Laura Tavanti, Marie Vermant, Arjun Nair, Iain Stewart, Sam M. Janes, Alexandra L. Young, David Barber, Daniel C. Alexander, Joanna C. Porter, Athol U. Wells, Mark G. Jones, Wim A. Wuyts, Joseph Jacob

## Abstract

**Background:** Idiopathic pulmonary fibrosis (IPF) with co-existent emphysema, termed combined pulmonary fibrosis and emphysema (CPFE) may be associated with reduced FVC decline compared to non-CPFE IPF patients. We examined associations between mortality and functional measures of disease progression in two IPF cohorts.

**Methods:** Visual emphysema extent (CPFE:non-CPFE: derivation cohort=317:183; replication cohort=358:152), scored on computed tomography imaging subgrouped CPFE patients using either a) 10%, or b) 15% visual emphysema threshold, or c) an unsupervised machine learning model considering emphysema and ILD extents. Baseline characteristics, 1-year forced vital capacity (FVC) and diffusion capacity for carbon monoxide (DLco) decline (linear mixed effects models), and their associations with mortality (multivariable Cox regression models) were compared across CPFE and non-CPFE subgroups.

**Results:** In both IPF cohorts, CPFE patients with >10% emphysema had a greater smoking history and lower baseline DLco compared to CPFE patients with <10% emphysema. Using multivariable Cox regression analyses in patients with >10% emphysema, 1-year DLco decline was a better indicator of mortality than 1-year FVC decline. Results were maintained in patients suitable for therapeutic IPF trials.

Results were replicated in the >15% emphysema population and using unsupervised machine learning. Importantly, the unsupervised machine learning approach identified CPFE patients in whom FVC decline did not associate strongly with mortality. In non-CPFE IPF patients, 1-year FVC declines >5% and >10% showed comparable mortality associations.

**Conclusion:** When assessing disease progression in IPF, DLco decline should be considered in patients with >10% emphysema and a >5% 1-year FVC decline threshold considered in non-CPFE IPF patients.

## Introduction

Emphysema is a common pulmonary finding on computed tomography (CT) imaging of idiopathic pulmonary fibrosis (IPF) patients [1]. The term combined pulmonary fibrosis and emphysema (CPFE) describes a potential clinical endotype characterized by the coexistence of upper lobe-predominant emphysema, lower lobe-predominant fibrosis and relative preservation of forced vital capacity (FVC) in the context of a disproportionately reduced gas transfer (DLco) [1–3]. CPFE is highly heterogeneous in terms of the distribution and relative extents of fibrosis and emphysema seen on CT.

CPFE patients are typically categorised using visual thresholds of emphysema extent: >0%, >5%, >10%, >15%. It has been suggested that a subset of CPFE patients (>15% emphysema) may manifest slower rates of FVC decline than CPFE patients with lesser amounts of emphysema [4]. Despite the importance of fibrosis in driving FVC decline, fibrosis extent hasn’t been considered in prior definitions of CPFE [5]. Categorisation of CPFE patients using a combination of fibrosis and emphysema is possible using data-driven machine learning methods. SuStaIn [6] is a machine learning method initially proposed for subtyping and modelling disease progression behaviour in dementia, which has been extended to COPD [7]. SuStaIn can identify disease subtypes with different progression patterns and can reconstruct their progression trajectories from cross-sectional data. A by-product of this approach would be the identification of patients in different CPFE subtypes who may benefit from different forms of disease progression monitoring, which in turn could inform clinical trial design.

In our study, we therefore aimed to assess whether FVC decline, the most widely used surrogate for mortality prediction in IPF associated with mortality in independent CPFE populations with >10% and >15% emphysema scored visually on CT imaging, and in CPFE subgroups categorised by considering relative extents of interstitial lung disease (ILD) and emphysema. We further examined whether DLco decline could represent an alternative surrogate for mortality in IPF patients with CPFE [5, 8].

## Methods

### Cohorts

Two independent IPF cohorts diagnosed by multidisciplinary teams were studied. The derivation cohort (n=500) derived from three centres: Ege University Hospital, Izmir, Turkey; St Antonius Hospital, Nieuwegein, Netherlands; Pisa University Hospital, Italy. The replication cohort (n=510) derived from four centres: University Hospital Southampton NHS Foundation Trust, UK; University College London Hospitals NHS Foundation Trust, UK; University Hospitals Leuven, Belgium; Australian IPF registry, Australia. Approval for this retrospective study of clinically indicated pulmonary function and CT data were obtained from the local research ethics committees and Leeds East Research Ethics Committee: 20/YH/0120.

### Visual CT Scoring of Emphysema and ILD

Patients with infection or cancer on baseline CT or who died within 3 months of the baseline CT were excluded from the study. Emphysema extent and fibrosis extent were visually scored in 6 lobes (the lingula was counted as the sixth lobe) by an experienced radiologist (JJ). Fibrosis extent comprised the sum of ground glass density (with overlying reticulation or traction bronchiectasis), reticulation, traction bronchiectasis and honeycomb cysts. Lobar extents of emphysema/fibrosis were summed and divided by 6 to obtain a lung percentage of emphysema/fibrosis. CPFE patients were subdivided in a primary analysis into those >10% emphysema (Figure 1), and in a secondary analysis into those >15% emphysema. A subset of 122 cases were evaluated independently by two radiologists (GC and JB: 3 and 4 years imaging experience respectively) to provide an estimate of observer variation in CT scores.

**Figure 1.**
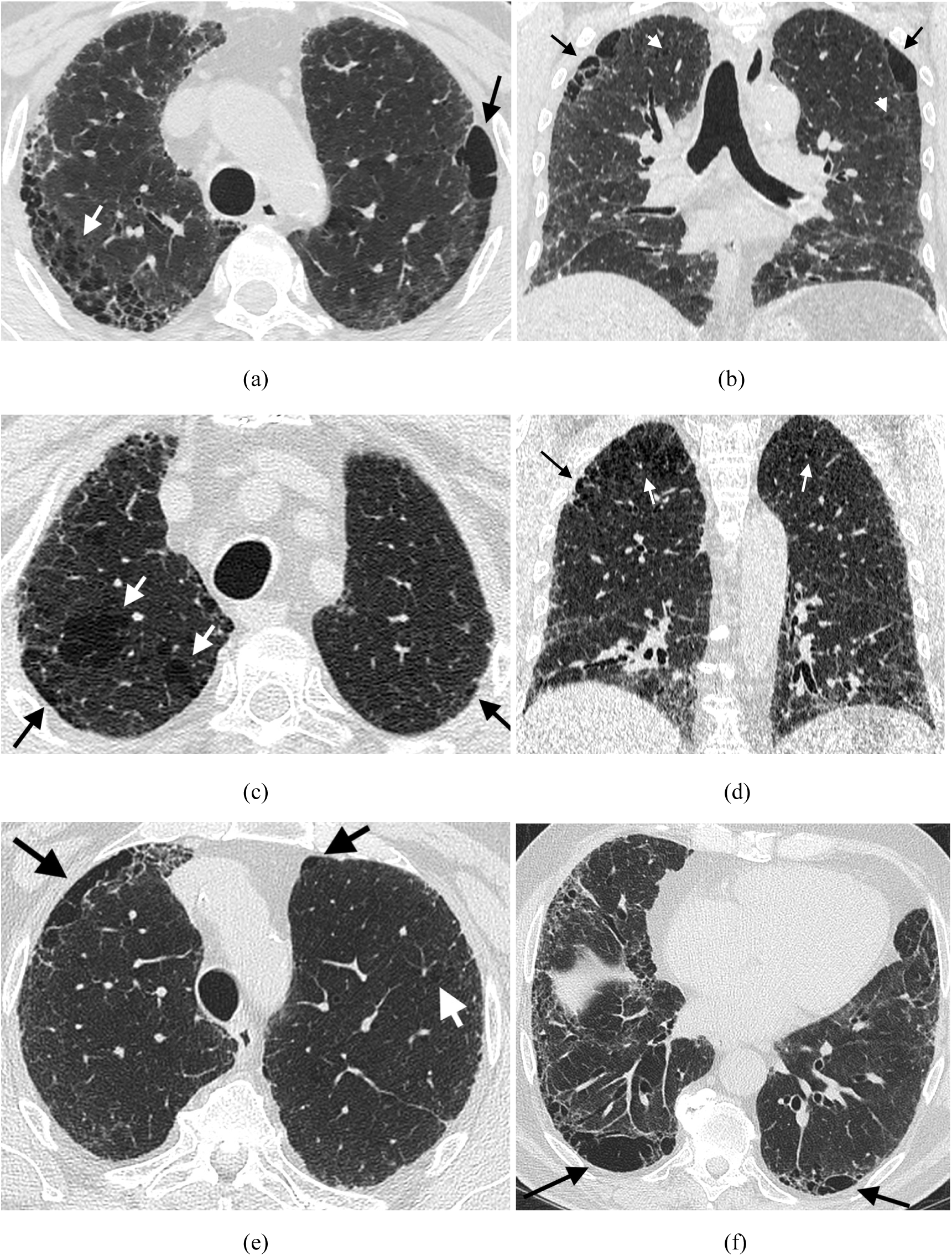
Computed tomography images of three subjects with 10% emphysema scored visually. A male 5-pack-year ex-smoker (age range: 56-60 years old) with axial (a) and coronal (b) imaging shows extensive upper lobe paraseptal emphysema (black arrows) and also centrilobular emphysema (white arrows) in a predominantly upper lobe distribution. Fibrosis with traction bronchiectasis, ground glass opacification and reticulation is seen in a lower zone predominant distribution. Figure c+d show respectively axial and coronal images of mixed paraseptal (black arrows) and centrilobular emphysema (white arrows) in a male 17-pack-year ex-smoker (age range: 56-60 years old). Axial images in a male 20-pack-year ex-smoker (age range: 71-75 years old) demonstrates a predominantly paraseptal distribution of emphysema (black arrows) in the upper (e) and lower (f) lobes with minimal centrilobular emphysema (white arrow).

### FVC/DLco Decline Modelling

Linear mixed-effects (LME) models estimated absolute and relative 1-year FVC decline and 1-year DLco decline. The trajectory of FVC for patients from different countries/centres was modelled separately by using the LME model. Fixed effects included: age at baseline CT date, gender, smoking history (never vs. ever), antifibrotics (never vs. ever), baseline percent predicted FVC (nearest to and within 3 months of baseline CT date), and time since baseline CT imaging date. Each subject had a random intercept and random slope. FVC measurements between baseline FVC date and 18 months after baseline CT date were used to build the LME model. Patients required at least two FVC measurements during this period and an FVC measurement within 3 months of baseline CT for study inclusion. Absolute and relative 1-year FVC declines were calculated. For relative 1-year FVC decline, each follow-up FVC measurement (mls) was divided by baseline FVC (mls) and multiplied by 100 [9] and LME-predicted relative FVC percentage calculated at 1 year. 1-year DLco decline was estimated using similar methods, with longitudinal DLco and baseline percent predicted DLco used in the LME models. LME models were implemented with MATLAB (version R2019b, Mathworks, Natick, Massachusetts, US).

### Machine learning delineation of CPFE subtypes

Only patients with emphysema scored visually in any lobe were considered for SuStaIn CPFE analysis. Using baseline data alone, SuStaIn can identify disease subtypes with distinct progression trajectories that describe the z-score evolution of multiple biomarkers. Z-scores for fibrosis and emphysema were calculated separately and were based on the interobserver variability (measured using the single determination standard deviation) between two radiologists visually estimating fibrosis and emphysema extent. For an individual CPFE subtype, fibrosis and emphysema within each of the six lobes was modelled as a monotonically increasing piece-wise linear function [6, 7]. The trained SuStaIn model, by reconstructing disease progression trajectories of each subtype, can predict probabilities that an individual belongs to a particular subtype and stage [6].

## Statistical analysis

Data are presented as means and standard deviations unless otherwise stated. Two-sample t-tests were used for continuous variables, and chi-squared tests were used for categorical variables. Kaplan-Meier survival plots and the log-rank test were used to test for differences in survival between non-CPFE IPF patients, and CPFE patients in different subgroups (using emphysema thresholds or SuStaIn subtype) in both IPF cohorts. Subanalyses were performed for patients satisfying lung function criterion for inclusion into IPF therapeutic trials (percent predicted DLco >30%, percent predicted FVC >50%, and forced expiratory volume in the first second/FVC ratio >0.7).

In multivariable mixed-effects Cox regression models associations of FVC decline and DLco decline with mortality were examined across IPF subtypes. Models were adjusted for age, gender, smoking history (never vs. ever), antifibrotic use (never vs. ever), and baseline disease severity (using percent predicted DLco at baseline). Differences between different countries/centres in each cohort were modelled by assigning a random intercept for each centre. Cox models were used with a minimum of 8 outcome events per predictor covariant [10]. The Concordance index (C-index) compared the goodness of fit of Cox regression models. P-values <0.01 were considered statistically significant. All mixed-effects Cox regression analyses were implemented by R (version 4.0.3 with Rstudio version 1.3.1093, Rstudio, Boston, Massachusetts, US).

To investigate the impact of emphysema on FVC and DLco decline in the different IPF subgroups (non-CPFE patients; CPFE patients classified using emphysema thresholds or SuStaIn), proportions of patients with >5% and >10% relative FVC decline in 1-year and >10% and >15% relative DLco decline in 1-year were calculated. Mean absolute 1-year FVC decline (mls) and DLco decline (mls) was also calculated for the three subgroups. Analyses were performed in both IPF cohorts, with subanalyses in subjects fulfilling criteria for inclusion into IPF therapeutic trials. Chi-squared tests with Bonferroni-adjusted p-values were calculated for categorical variables. A one-way ANOVA test examined differences in mean absolute FVC decline (ml/year) with a post hoc Tukey Honest Significant Difference (HSD) test used to compare pairwise differences in subtypes.

## Results

### Baseline characteristics

317/500 (63%) IPF patients in the derivation cohort had emphysema and were defined as CPFE compared to 358/510 (70%) IPF patients with CPFE in the replication cohort. CPFE patients were more likely to be smokers, had a higher percent-predicted FVC and lower percent-predicted DLco than non-CPFE patients.

Across the derivation and replication cohorts, CPFE patients with >10% emphysema comprised greater numbers of smokers and had lower baseline percent predicted DLco compared to CPFE patients with <10% emphysema (Table 1). To power analyses, patients in both IPF cohorts fulfilling entry criteria for therapeutic trials were combined into a single cohort (Supplementary Table 1). Baseline characteristics of CPFE patients with emphysema above or below 15% in derivation and replication cohorts are shown in Supplementary Tables 2-3.

**Table 1.** Baseline characteristics of non-CPFE IPF patients and CPFE patients with emphysema below or above 10% in the derivation and replication cohorts.

| Cohort | Variable | Non-CPFE IPF patients | CPFE patients with emphysema < 10% | CPFE patients with emphysema ≥ 10% | P-value |
| --- | --- | --- | --- | --- | --- |
| Derivation cohort | Subjects (%) | 183 (36.6) | 174 (34.8) | 143 (28.6) | - |
|  | Age (years) | 67.8±9.2 | 66.9±9.1 | 65.0±9.1 | 0.06 |
|  | Male (%) | 110/183 (60.1) | 143/174 (82.2) | 132/143 (92.3) | 0.01 |
|  | Never-/ever-smokers (ever %) | 92/91 (49.7) | 38/133 (77.8) * | 8/134 (94.4) ** | < 0.0001 |
|  | Visual fibrosis extent (%) | 38.7±14.6 | 36.3±14.1 | 40.8±13.5 | 0.004 |
|  | Visual emphysema extent (%) | 0±0 | 4.76±2.3 | 20.4±8.8 | < 0.0001 |
|  | FVC (% predicted, n) | 77.1±20.8 (158) | 80.1±20.2 (150) | 79.1±21.9 (122) | 0.68 |
|  | DLco (% predicted, n) | 52.2±16.5 (151) | 51.6±15.1 (138) | 40.4±13.3 (116) | < 0.0001 |
| Replication cohort | Subjects (%) | 152 (29.8) | 206 (40.4) | 152 (29.8) | - |
|  | Age (years) | 71.6±8.4 | 71.9±8.3 | 70.5±8.0 | 0.12 |
|  | Male (%) | 96/152 (63.2) | 168/206 (81.6) | 128/152 (84.2) | 0.60 |
|  | Never-/ever-smokers (ever %) | 78/74 (48.7) | 51/152 (74.9) † | 22/129 (85.4) †† | 0.02 |
|  | Visual fibrosis extent (%) | 34.0±14.9 | 34.6±12.8 | 37.8±12.4 | 0.02 |
|  | Visual emphysema extent (%) | 0±0 | 4.9±2.4 | 21.1±11.1 | < 0.0001 |
|  | FVC (% predicted, n) | 84.5±21.1 (137) | 84.4±20.5 (184) | 86.6±18.9 (137) | 0.32 |
|  | DLco (% predicted, n) | 55.2±15.1 (121) | 51.2±16.0 (176) | 40.7±11.2 (126) | < 0.0001 |
| FVC: forced vital capacity; DLco: diffusing capacity for carbon monoxide; * 171 patients and ** 142 patients had smoking data available in derivation cohort; † 203 patients and †† 151 patients had smoking data available in replication cohort; P-value shows the significance of the difference between CPFE patients with emphysema above or below 10%. |  |  |  |  |  |

The interobserver variation in visual emphysema scores, measured using Cohens Kappa for 0%, 5%, 10%, and 15% emphysema thresholds was: 0.2, 0.5, 0.61, 0.69, respectively demonstrating substantial agreement for a 10% visual emphysema threshold.

### Machine Learning Model

Machine learning analyses of ILD and emphysema extents in the CPFE population identified two distinct CPFE subtypes. One subtype (*Fibrosis Dominant CPFE*; 60% of derivation cohort CPFE patients and 61% of replication cohort CPFE patients) had much more extensive fibrosis at an early stage followed by a later emergence of emphysema (Supplementary Figure 3 and 4). The second subtype (*Matched CPFE*) demonstrated fibrosis and emphysema worsening together, with later stages showing relatively more extensive emphysema and less fibrosis compared to the fibrosis-dominant CPFE subtype (Supplementary Table 4 and 5).

### PFT decline analyses

Fewer CPFE patients with >10% emphysema reached the >10% or >5% 1-year FVC decline thresholds and had lower mean absolute FVC declines, though differences between groups did not reach statistical significance (Table 2). Greater numbers of CPFE patients with >10% emphysema demonstrated 1-year DLco declines >15%, though again results did not reach statistical significance (Table 3). Similar trends were found in the replication cohort, patients fulfilling criteria to enter IPF therapeutic trials (Table 2 and 3), and when CPFE was categorized using a 15% emphysema threshold or machine learning analyses (Supplementary Table 6 and 7).

**Table 2.** FVC decline analysis in different subgroups of IPF patients

| Cohort | Subgroup | FVC data available<br>cases/all case | Relative 1-year FVC decline (%) |  | Absolute 1-year FVC<br>decline<br>(mls/year) |
| --- | --- | --- | --- | --- | --- |
|  |  |  | Number of >10%<br>(proportion) | Number of >5%<br>(proportion) | Mean |
| Derivation<br>cohort | Non-CPFE | 150/183 | 51 (34%) | 81 (54%) | 163.50 |
|  | CPFE with emphysema <10% | 136/174 | 39 (28.68%) | 69 (50.74%) | 180.12 |
|  | CPFE with emphysema ≥10% | 115/143 | 27 (23.48%) | 49 (42.61%) | 97.43 |
| Replication<br>cohort | Non-CPFE | 124/152 | 24 (19.35%) | 50 (40.32%) | 110.65 |
|  | CPFE with emphysema <10% | 170/206 | 37 (21.76%) | 75 (44.12%) | 132.62 |
|  | CPFE with emphysema ≥10% | 130/152 | 21 (16.15%) | 44 (33.85%) | 87.71 |
| Combined<br>drug trial<br>cohort | Non-CPFE | 222/236 | 59 (26.58%) | 105 (47.30%) | 142.94 |
|  | CPFE with emphysema <10% | 240/261 | 57 (23.75%) | 113 (47.08%) | 164.81 |
|  | CPFE with emphysema ≥10% | 150/157 | 29 (19.33%) | 56 (37.33%) | 112.19 |

The proportions of patients with more than 10% and 5% relative 1-year FVC decline, and the mean of absolute 1-year FVC decline in derivation, replication cohorts and combined drug trial cohort (patients fulfilling criteria to enter IPF therapeutic trials in derivation and replication cohorts) are shown in this table. The number of subjects with available FVC decline versus the number of all subjects belonging to a certain subgroup is shown in n/n format. We also compared a) non-CPFE with CPFE with emphysema ≥10%, b) CPFE with emphysema ≥10% and CPFE with emphysema <10%, in terms of the relative decline and absolute decline. None of the results were statistically significantly different. CPFE: combined pulmonary fibrosis and emphysema; IPF: idiopathic pulmonary fibrosis; FVC: forced vital capacity.

**Table 3.** DLco decline analysis in different subgroups of IPF patients

| Cohort | Subgroup | DLco data available cases/all case | Relative 1-year DLco decline (%) |  | Absolute 1-year DLco decline (mls/year) |
| --- | --- | --- | --- | --- | --- |
|  |  |  | Number of >15% (proportion) | Number of >10% (proportion) | Mean |
| Derivation cohort | Non-CPFE | 132/183 | 52 (39.39%) | 73 (55.30%) | 645.39 |
|  | CPFE with emphysema <10% | 125/174 | 42 (33.60%) | 60 (48%) | 1020.97 |
|  | CPFE with emphysema ≥10% | 107/143 | 42 (39.25%) | 59 (55.14%) | 870.88 |
| Replication cohort | Non-CPFE | 108/152 | 30 (27.78%) | 43 (39.81%) | 769.10 |
|  | CPFE with emphysema <10% | 161/206 | 38 (23.60%) | 67 (41.61%) | 615.04 |
|  | CPFE with emphysema ≥10% | 117/152 | 42 (35.90%) | 64 (54.70%) | 581.21 |
| Combined drug trial cohort | Non-CPFE | 213/236 | 71 (33.33%) | 100 (46.95%) | 748.91 |
|  | CPFE with emphysema <10% | 238/261 | 66 (27.73%) | 112 (47.06%) | 863.75 |
|  | CPFE with emphysema ≥10% | 146/157 | 54 (36.99%) | 80 (54.79%) | 814.72 |

The proportions of patients with more than 15% and 10% relative 1-year DLco decline, and the mean of absolute 1-year DLco decline in derivation, replication cohorts and combined drug trial cohort (patients fulfilling criteria to enter IPF therapeutic trials in derivation and replication cohorts) are shown in this table. The number of subjects with available DLco decline versus the number of all subjects belonging to a certain subgroup is shown in n/n format. We also compared a) non-CPFE with CPFE with emphysema ≥10%, b) CPFE with emphysema ≥10% and CPFE with emphysema <10%, in terms of the relative decline and absolute decline. None of the results were statistically significantly different. CPFE: combined pulmonary fibrosis and emphysema; IPF: idiopathic pulmonary fibrosis; DLco: diffusion capacity for carbon monoxide.

### Survival Analyses

Kaplan-Meier survival plots (Figure 2) demonstrated that in both cohorts, non-CPFE and CPFE patients with <10% emphysema had a significantly better prognosis than CPFE patients with >10% emphysema. Results were maintained in patients fulfilling criteria to enter IPF therapeutic trials and were similar when CPFE patients were separated using a 15% emphysema threshold or machine learning analyses (Supplementary Figure 1 and 2).

**Figure 2.**
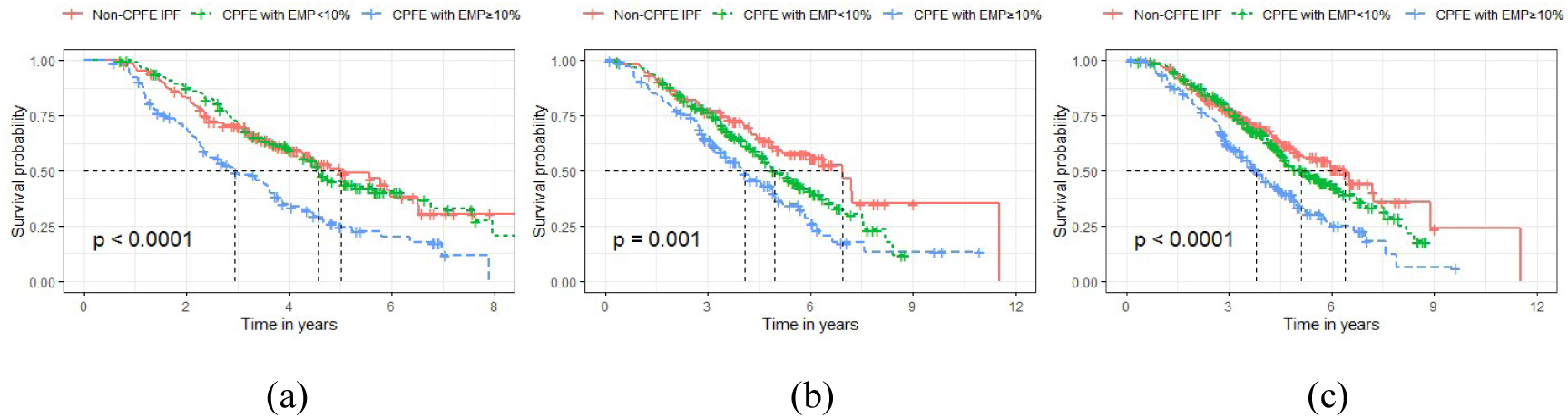
Kaplan-Meier curves of non-CPFE IPF patients (red), CPFE patients with emphysema <10% (green) and CPFE patients with emphysema ≥ 10% (blue) in the derivation cohort (a), the replication cohort (b), combined derivation and replication cohort patients qualifying for therapeutic trials (c). Log-rank tests show a significant difference in mortality between the three subtypes in all three analyses.

### Mortality analysis for visual emphysema thresholds

Multivariable Cox regression models adjusted for patient age, gender, smoking history (never vs. ever), antifibrotic use (never vs. ever), and baseline percent predicted DLco showed that across both study cohorts, in non-CPFE patients, a 5% 1-year FVC decline threshold showed equivalent associations with mortality as compared to a 10% 1-year FVC decline threshold (Tables 4 and 5). A 5% 1-year FVC decline threshold identified more non-CPFE patients (derivation cohort=59%; replication cohort=108%) than a 10% 1-year FVC decline threshold (Table 2). Associations with mortality were maintained in patients fulfilling criteria to enter IPF therapeutic trials (Supplementary Table 8), where 78% more non-CPFE patients had >5% 1-year FVC declines compared to patients with >10% 1-year FVC decline (Table 2).

**Table 4.**
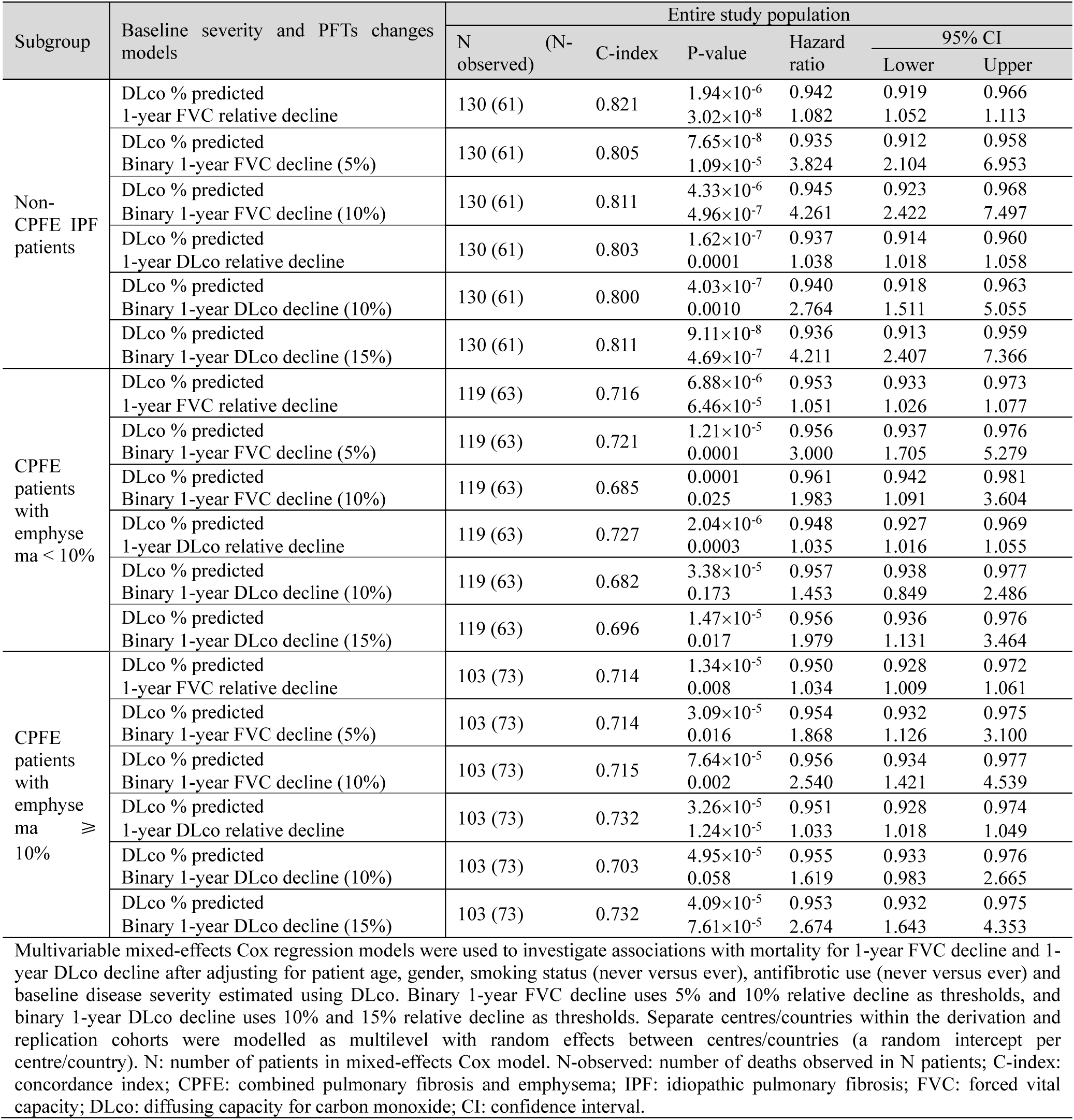
Multivariable mixed-effects Cox proportional hazards regression models in non-CPFE patients and the two CPFE subgroups in the derivation IPF cohorts.

**Table 5.**
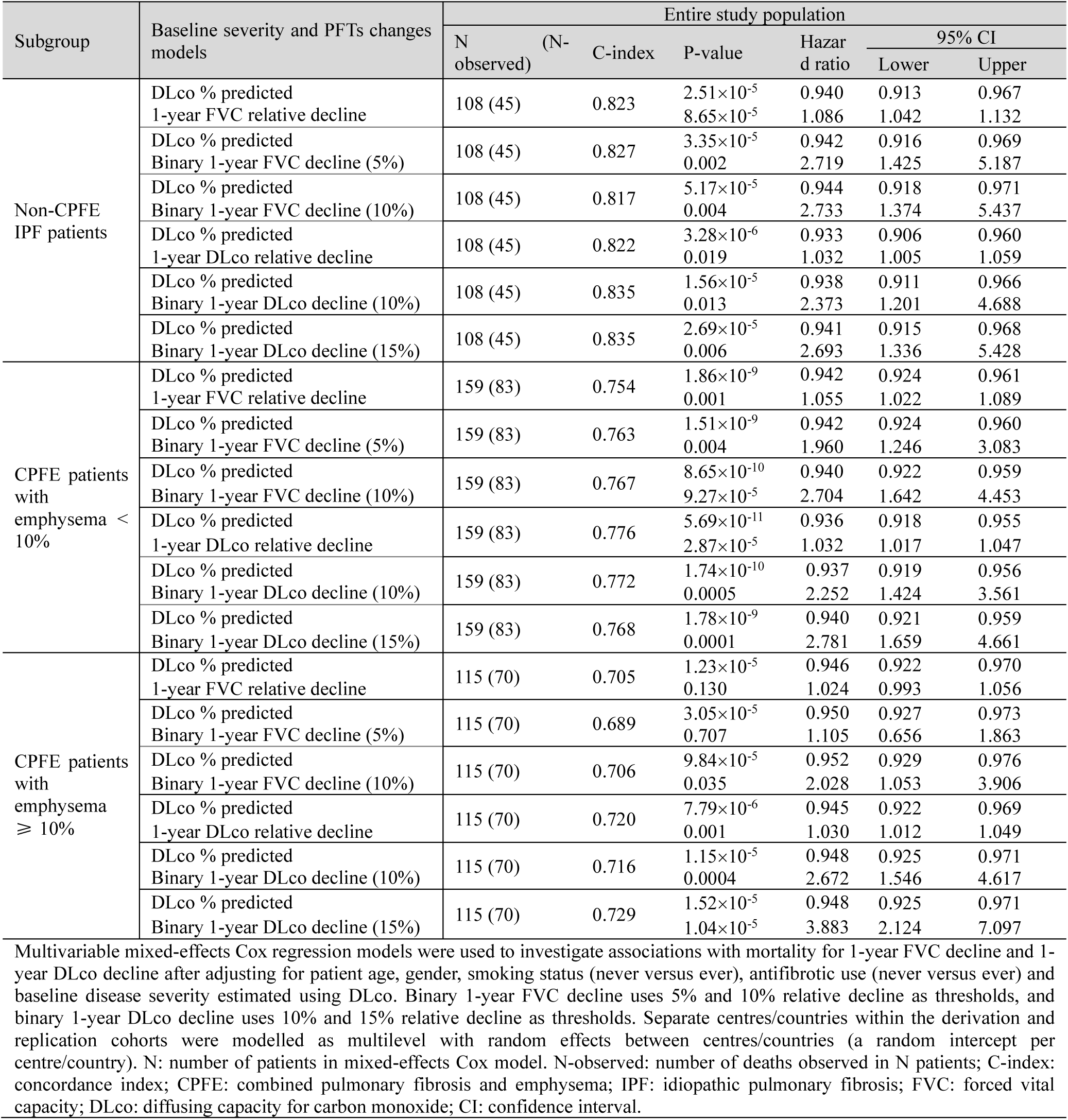
Multivariable mixed-effects Cox proportional hazards regression models in non-CPFE patients and the two CPFE subgroups in the replication IPF cohorts.

For CPFE patients with >10% emphysema (derivation cohort n=103/352 (29%); replication cohort n=115/382 (30%)), 1-year DLco decline showed a much stronger association with mortality than 1-year FVC decline in derivation and replication cohorts (Table 4 and 5). 1-year FVC decline did not associate significantly with mortality in the replication cohort (Table 5). When DLco thresholds were examined in CPFE patients with >10% emphysema in both cohorts, >15% 1-year DLco decline showed stronger associations with mortality than >10% 1-year FVC decline. In subjects eligible for inclusion into IPF therapeutic trials (where 144/589 (24%) patients had >10% emphysema) DLco decline showed stronger associations with mortality than FVC decline (Supplementary Table 8). Similar trends were observed in multivariable analyses performed in CPFE patients with >15% emphysema (Supplementary Table 9-11).

### Mortality analyses of machine learning derived CPFE subgroups

Trends seen for the 10% visual emphysema threshold were again replicated when CPFE patients were separated using machine learning analyses that considered ILD and emphysema extents. The *Matched CPFE* cohort better delineated patients in whom FVC decline proved a poor surrogate for mortality compared to visual emphysema thresholds of >10% and >15%. Importantly, in the *Matched CPFE* cohort, DLco decline, whether measured as absolute decline or a >15% DLco threshold remained a strong surrogate for mortality (Supplementary Table 12-14).

## Discussion

Our study evaluated functional indicators of disease progression in IPF patients with emphysema that have been the key mortality surrogates used in clinical care and therapeutic trials. We identified three important findings across two IPF populations: Firstly, we demonstrated the limited associations between FVC decline and mortality in CPFE patients with >10% and >15% emphysema, and conversely the strong associations with mortality for DLco decline in the same subgroups. Second, our machine learning model identified a subgroup of CPFE patients where a relatively greater amount of emphysema compared to ILD accentuated the limited associations between ILD-driven FVC decline and mortality in these CPFE patients. Lastly, in non-CPFE patients we showed that FVC decline is a powerful measure of IPF progression showing comparable associations with mortality for both >5% and >10% 1-year FVC decline thresholds. Using a >5% 1-year FVC decline threshold in non-CPFE patients identified over 50% more subjects with real declines than when using a >10% 1-year FVC decline threshold.

FVC decline occupies a cardinal role in the assessment of disease progression in IPF as it has been shown to be a strong surrogate for mortality [11]. The demonstration however that FVC decline may be curtailed in IPF patients with >15% [4] emphysema raised the question of whether FVC decline remained a surrogate for mortality in IPF patients with more extensive emphysema. Only one other study, by Schmidt et al [8], which was relatively underpowered (n=42) for subjects with moderate/severe emphysema (defined as emphysema at least as extensive as ILD), addressed this question and found that FVC decline did not associate with mortality at 12 months. Other studies considering IPF patients regardless of emphysema presence/extent have shown strong associations between mortality and other functional decline measures/thresholds including: DLco decline thresholds of >10% [12] and 15% [13], and FVC declines of >5% [14–16].

An explanation for the poor association between FVC decline and mortality in patients with more extensive emphysema may relate to the impact of fibrosis when encroaching on areas of emphysema. Emphysematous regions of lung commonly demonstrate air trapping as thickened small airways collapse on expiration. Fibrotic processes however can irreversibly pull open small airways. The supervening traction bronchiolectasis can result in emphysematous airspaces being ventilated, thereby artificially preserving FVC. In IPF patients with emphysema, as fibrosis progresses and extends to involve the upper zones of the lungs, more emphysematous lung may become incorporated into the expiratory lung volume over time. A consequence may more heterogeneity in expiratory volumes superimposing considerable noise to the overarching pattern of progressive FVC decline. This effect is likely to be more pronounced in patients with more extensive emphysema.

One limitation in prior definitions of CPFE has been the focus on emphysema extent alone as the sole arbiter for categorising a CPFE endotype. A recent ATS/ERS/ALAT/JRS research statement identified a 5% emphysema threshold as a research definition for CPFE patients, whilst suggesting a 15% emphysema threshold for classifying a CPFE clinical syndrome [5]. In our study we found that a 10% emphysema threshold (which showed substantial CT observer agreement) may represent a better cut-off than a 15% emphysema threshold to identify a CPFE population disenfranchised by the use of FVC as a sole measure of disease progression.

A further challenge with CPFE definitions being determined by emphysema thresholds is that FVC decline is primarily driven by ILD progression rather than emphysema progression. Our novel unsupervised machine learning model (SuStaIn) considered both fibrosis and emphysema when subtyping patients and replicated the strong association of DLco decline and mortality in patients with more extensive emphysema seen in CPFE patients with >10% emphysema. By considering ILD extent in relation to emphysema extent, the SuStaIn model improved delineation of a subgroup of CPFE patients, fulfilling criteria to enter IPF therapeutic trials, where FVC decline did not associate strongly with mortality. This could have implications for assessing disease progression in future IPF clinical trials.

Prior studies have shown associations between DLco decline and mortality in IPF [8, 12, 13, 17–19] but have not analysed the impact of emphysema on DLco trends. DLco decline has generally been less consistent in its links with mortality than FVC decline in IPF patients [20]. Yet DLco decline may have particular relevance in subsets of IPF patients [21]. For example, the strong mortality signal for DLco decline seen in CPFE patients with more extensive emphysema could reflect progressive localised pulmonary hypertension complicating CPFE patients with more extensive emphysema [22, 23].

There were limitations to the current study. A single observer scored the CTs for fibrosis and emphysema. For studies to be clinically meaningful, they have to be suitably powered, and this requires the careful evaluation of large IPF populations which is challenging with limited availability of radiologists. The single read of CTs in this study aligns with other large scale IPF studies where pragmatic considerations required assessment of CTs by a single specialist [24, 25]. Similar functional measures and IPF subgroups proportions across both study cohorts provides reassurance for the validity of the visual CT scores. The improvement in observer agreement at higher emphysema thresholds (even amongst less experienced radiologists) adds confidence to the reliability of visual scores at an emphysema threshold of 10%. This also aligns with prior work [26] demonstrating improved interobserver agreement at emphysema extent categories of 10% and 15% versus 0% and 5%. Lastly, whilst we would have liked to have fully automated our machine learning model, using computationally quantified emphysema as an objective measure of disease, no existing automated tools can reliably distinguish emphysema from honeycombing and traction bronchiectasis. Accordingly, there will remain a reliance on visual CT reads for assessing the emphysematous component of CPFE in the near future.

In conclusion, annual DLco decline was shown to be a better mortality surrogate for patients with more than 10% emphysema than FVC decline. Findings were validated by a data-driven machine learning method that considers emphysema and ILD extents when defining patients with more extensive emphysema. These observations may be useful in clinical trial design to identify subjects where FVC decline is a poor disease progression measure. A 5% 1-year FVC decline threshold however was found to be a comparable mortality indicator to a 1-year 10% FVC decline threshold in non-CPFE IPF patients.

## Data Availability

Approvals are not in place to share the data produced in the present study.

## Acknowledgments

This research was funded in whole or in part by the Wellcome Trust [209553/Z/17/Z]. For the purpose of open access, the author has applied a CC-BY public copyright licence to any author accepted manuscript version arising from this submission. This project, JJ, EG, ED, SMJ and JCP were also supported by the NIHR UCLH Biomedical Research Centre, UK. MGJ, TJMW and CJB acknowledge the support of the NIHR Southampton Biomedical Research Centre. AZ was supported by CSC-UCL Joint Research Scholarship. The Australian IPF Registry is an initiative of Lung Foundation Australia and is supported by Foundation partners Boehringer Ingelheim, Roche Products Pty. Limited.

## Author Contributions

AZ, EG, IS, ALY, DB, DCA, AUW, and JJ contributed to study design and data interpretation. AZ, EG, NM, MGJ, CvM, TJC, CR, RC, TJMW, ED, TG, RS, AA, CJB, HWvE, HJ, ADL, MD, KP, LJDS, FvB, JB, GC, AP, MV, PH, YM, AT, MT, SV, LT, MV, AN, SMJ, JCP, MGJ, WAW and JJ were responsible for data acquisition. AZ, EG, IS, and JJ contributed to the statistical analysis. AZ and JJ prepared the first draft of the manuscript. AZ and JJ were responsible for study data integrity. All authors reviewed the manuscript and approved the final submitted version.

