## Supplementary for "Mortality surrogates in combined pulmonary fibrosis and emphysema"

### Supplementary Appendix

Supplementary Table 1. Baseline characteristics of non-CPFE IPF patients and CPFE patients fulfilling criteria to enter IPF therapeutic trials and with emphysema below or above 10% in the combined cohorts.

| Variable | Non-CPFE IPF patients | CPFE patients with emphysema < 10% | CPFE patients with emphysema ⩾ 10% | P-value |
|---|---|---|---|---|
| Subjects (%) | 236 (36.1) | 261 (39.9) | 157 (24.0) | - |
| Age (years) | 69.8±8.2 | 69.8±8.8 | 67.5±9.0 | 0.009 |
| Male (%) | 141/236 (59.7) | 209/261 (80.1) | 142/157 (90.4) | 0.008 |
| Never-/ever-smokers (ever %) | 121/115 (48.7) | 64/192 (75) * | 15/140 (90.3) ** | 0.0002 |
| Visual fibrosis extent (%) | 34.4±13.9 | 33.9±12.9 | 37.5±13.0 | 0.007 |
| Visual emphysema extent (%) | 0±0 | 4.7±2.3 | 18.7±8.4 | < 0.0001 |
| FVC (% predicted) | 83.9±19.1 | 85.2±17.7 | 85.7±17.9 | 0.81 |
| DLco (% predicted) | 55.7±14.1 | 53.1±13.7 | 45.7±9.8 | < 0.0001 |

FVC: forced vital capacity; DLco: diffusing capacity for carbon monoxide; * 256 patients and ** 155 patients had smoking data available. P-value shows the significance of the difference between CPFE patients with emphysema above or below 10%.

Supplementary Table 2. Baseline characteristics of non-CPFE IPF patients and CPFE patients with emphysema below or above 15% in the derivation and replication cohorts.

| Cohort | Variable | Non-CPFE IPF patients | CPFE patients with emphysema < 15% | CPFE patients with emphysema ⩾ 15% | P-value |
|---|---|---|---|---|---|
| Derivation cohort | Subjects (%) | 183 (36.6) | 218 (43.6) | 99 (19.8) | - |
| | Age (years) | 67.8±9.2 | 66.3±9.1 | 65.4±9.2 | 0.39 |
| | Male (%) | 110/183 (60.1) | 185/218 (84.9) | 90/99 (90.9) | 0.20 |
| | Never-/ever-smokers (ever %) | 92/91 (49.7) | 40/174 (81.3) * | 6/93 (93.9) | 0.006 |
| | Visual fibrosis extent (%) | 38.7±14.6 | 37.3±13.9 | 40.8±14.0 | 0.04 |
| | Visual emphysema extent (%) | 0±0 | 6.2±3.6 | 24.2±8.2 | < 0.0001 |
| | FVC (% predicted, n) | 77.1±20.8 (158) | 78.7±20.4 (189) | 81.7±22.1 (83) | 0.30 |
| | DLco (% predicted, n) | 52.2±16.5 (151) | 50.1±14.7 (174) | 38.7±13.9 (80) | < 0.0001 |
| Replication cohort | Subjects (%) | 152 (29.8) | 258 (50.59) | 100 (19.6) | - |
| | Age (years) | 71.6±8.4 | 71.7±8.1 | 70.3±8.6 | 0.17 |
| | Male (%) | 96/152 (63.2) | 211/258 (81.8) | 85/100 (85) | 0.57 |
| | Never-/ever-smokers (ever %) | 78/74 (48.7) | 60/195 (76.5) [†] | 13/86 (86.9) [††] | 0.04 |
| | Visual fibrosis extent (%) | 34.0±14.9 | 35.2±12.9 | 37.7±11.9 | 0.08 |
| | Visual emphysema extent (%) | 0±0 | 6.3±3.6 | 26.0±10.9 | < 0.0001 |
| | FVC (% predicted, n) | 84.5±21.1 (137) | 84.3±20.4 (227) | 87.8±18.3 (94) | 0.13 |
| | DLco (% predicted, n) | 55.2±15.1 (121) | 49.7±15.5 (215) | 39.6±11.4 (87) | < 0.0001 |

FVC: forced vital capacity; DLco: diffusing capacity for carbon monoxide; * 214 patients had smoking data available in the derivation cohort; [†] 255 patients and [††] 99 patients had smoking data available in the replication cohort. P-value shows the significance of the difference between CPFE patients with emphysema above or below 15%.

Supplementary Table 3. Baseline characteristics of non-CPFE IPF patients and CPFE patients fulfilling criteria to enter IPF therapeutic trials and with emphysema below or above 15% in the combined cohorts.

| Variable | Non-CPFE IPF patients | CPFE patients with emphysema < 15% | CPFE patients with emphysema ⩾ 15% | P-value |
|---|---|---|---|---|
| Subjects (%) | 236 (36.1) | 318 (48.6) | 100 (15.3) | - |
| Age (years) | 69.8±8.2 | 69.4±8.9 | 67.4±8.9 | 0.06 |
| Male (%) | 141/236 (59.7) | 260/318 (81.8) | 91/100 (91) | 0.04 |
| Never-/ever-smokers (ever %) | 121/115 (48.7) | 71/241 (77.2) * | 8/91 (91.9) ** | 0.002 |
| Visual fibrosis extent (%) | 34.4±13.9 | 35.0±13.1 | 36.1±12.8 | 0.43 |
| Visual emphysema extent (%) | 0±0 | 6.0±3.5 | 22.6±8.2 | < 0.0001 |
| FVC (% predicted) | 83.9±19.1 | 84.7±17.7 | 87.5±17.8 | 0.18 |
| DLco (% predicted) | 55.7±14.1 | 52.0±13.1 | 45.0±10.4 | < 0.0001 |

FVC: forced vital capacity; DLco: diffusing capacity for carbon monoxide; * 312 patients and ** 99 patients had smoking data available. P-value shows the significance of the difference between CPFE patients with emphysema above or below 15%.

Supplementary Table 4. Baseline characteristics of non-CPFE IPF patients and CPFE patients in the fibrosis-dominant and Matched-CPFE subtypes in the derivation and replication cohorts.

| Cohort | Variable | Non-CPFE IPF patients | Fibrosis-dominant CPFE subtype | Matched-CPFE subtype | P-value |
|---|---|---|---|---|---|
| Derivation cohort | Subjects (%) | 183 (36.6) | 191 (38.2) | 126 (25.2) | - |
| | Age (years) | 67.8±9.2 | 66.7±9.1 | 65.0±9.1 | 0.09 |
| | Male (%) | 110/183 (60.1) | 159/191 (83.2) | 116/126 (92.1) | 0.04 |
| | Never-/ever-smokers (ever %) | 92/91 (49.7) | 40/148 (78.7) * | 6/119 (95.2) ** | 0.0001 |
| | Visual fibrosis extent (%) | 38.7±14.6 | 38.6±14.2 | 38.1±13.7 | 0.77 |
| | Visual emphysema extent (%) | 0±0 | 5.6±3.4 | 21.3±9.1 | < 0.0001 |
| | FVC (% predicted, n) | 77.1±20.8 (158) | 78.3±19.9 (167) | 81.8±22.5 (105) | 0.19 |
| | DLco (% predicted, n) | 52.2±16.5 (151) | 50.2±15.4 (153) | 40.9±13.4 (101) | < 0.0001 |
| Replication cohort | Subjects (%) | 152 (29.8) | 227 (44.5) | 131 (25.7) | - |
| | Age (years) | 71.6±8.4 | 71.8±8.3 | 70.5±8.1 | 0.14 |
| | Male (%) | 96/152 (63.2) | 187/227 (82.4) | 109/131 (83.2) | 0.96 |
| | Never-/ever-smokers (ever %) | 78/74 (48.7) | 56/168 (75) † | 17/113 (86.9) †† | 0.01 |
| | Visual fibrosis extent (%) | 34.0±14.9 | 37.2±12.6 | 33.8±12.6 | 0.01 |
| | Visual emphysema extent (%) | 0±0 | 5.8±3.6 | 22.1±11.7 | < 0.0001 |
| | FVC (% predicted, n) | 84.5±21.1 (137) | 83.1±20.4 (200) | 88.9±18.4 (121) | 0.01 |
| | DLco (% predicted, n) | 55.2±15.1 (121) | 49.8±16.1 (189) | 41.8±11.7 (113) | < 0.0001 |

FVC: forced vital capacity; DLco: diffusing capacity for carbon monoxide; *188 patients and **125 patients had smoking data available in derivation cohort; †224 patients and ††130 patients had smoking data available in replication cohort. P-value shows the significance of the difference between CPFE patients in the fibrosis-dominant and Matched-CPFE subtypes.

Supplementary Table 5. Baseline characteristics of non-CPFE IPF patients and Fibrosis-dominant and Matched-CPFE subtypes fulfilling criteria to enter IPF therapeutic trials in the combined cohorts.

| Variable | Non-CPFE IPF patients | Fibrosis-dominant CPFE subtype | Matched-CPFE subtype | P-value |
|---|---|---|---|---|
| Subjects (%) | 236 (36.1) | 281 (43.0) | 137(20.9) | - |
| Age (years) | 69.8±8.2 | 69.6±8.9 | 67.5±8.8 | 0.02 |
| Male (%) | 141/236 (59.7) | 230/281 (81.9) | 121/137 (88.3) | 0.12 |
| Never-/ever-smokers (ever %) | 121/115 (48.7) | 66/210 (76.1) * | 13/122 (90.4) ** | 0.0009 |
| Visual fibrosis extent (%) | 34.4±13.9 | 36.5±13.1 | 32.6±12.6 | 0.004 |
| Visual emphysema extent (%) | 0±0 | 5.4±3.4 | 19.2±9.0 | < 0.0001 |
| FVC (% predicted) | 83.9±19.1 | 84.3±17.6 | 87.7±17.7 | 0.06 |
| DLco (% predicted) | 55.7±14.1 | 52.1±13.7 | 46.7±10.0 | < 0.0001 |

FVC: forced vital capacity; DLco: diffusing capacity for carbon monoxide; * 276 patients and ** 135 patients had smoking data available. P-value shows the significance of the difference between CPFE patients in the fibrosis-dominant and Matched-CPFE subtypes.

Supplementary Table 6. FVC decline analysis in different subgroups of IPF patients.

| Cohort | Subgroup | FVC data available cases/all case | Relative 1-year FVC decline (%) | | Absolute 1-year FVC decline (mls/year) |
|---|---|---|---|---|---|
| | | | Number of >10% (proportion) | Number of >5% (proportion) | Mean |
| Derivation cohort | Non-CPFE | 150/183 | 51 (34%) | 81 (54%) | 163.50 |
| | CPFE with emphysema <15% | 174/218 | 51 (29.31%) | 90 (51.72%) | 165.21 |
| | CPFE with emphysema ⩾15% | 77/99 | 15 (19.48%) | 28 (36.36%) | 90.31 |
| | Fibrosis-dominant CPFE | 153/191 | 46 (30.07%) | 77 (50.33%) | 159.50 |
| | Matched-CPFE | 98/126 | 20 (20.41%) | 41 (41.84%) | 115.27 |
| Replication cohort | Non-CPFE | 124/152 | 24 (19.35%) | 50 (40.32%) | 110.65 |
| | CPFE with emphysema <15% | 211/258 | 43 (20.38%) | 91 (43.13%) | 127.95 |
| | CPFE with emphysema ⩾15% | 89/100 | 15 (16.85%) | 28 (31.46%) | 78.10 |
| | Fibrosis-dominant CPFE | 187/227 | 41 (21.93%) | 83 (44.39%) | 135.32 |
| | Matched-CPFE | 113/131 | 17 (15.04%) | 36 (31.86%) | 76.48 |
| Combined drug trial cohort | Non-CPFE | 222/236 | 59 (26.58%) | 105 (47.30%) | 142.94 |
| | CPFE with emphysema <15% | 295/318 | 71 (24.07%) | 141 (47.80%)* | 161.88 |
| | CPFE with emphysema ⩾15% | 95/100 | 15 (15.79%) | 28 (29.47%)† | 90.84 |
| | Fibrosis-dominant CPFE | 262/281 | 65 (24.81%) | 124 (47.33%) | 163.21 |
| | Matched-CPFE | 128/137 | 21 (16.41%) | 45 (35.16%) | 106.42 |

The proportions of patients with more than 10% and 5% relative 1-year FVC decline, and the mean of absolute 1-year FVC decline in different subgroups in derivation and replication cohorts and the combined drug trial cohort (patients fulfilling criteria to enter IPF therapeutic trials in derivation and replication cohorts) are shown in this table. The number of subjects with available FVC decline versus the number of all subjects within a subgroup is shown in n/n format. We also compared a) non-CPFE with CPFE with emphysema ⩾15%, b) CPFE with emphysema ⩾15% and CPFE with emphysema <15%, c) non-CPFE with Matched-CPFE subtype, d) Fibrosis-dominant CPFE subtype and Matched-CPFE subtype in terms of the relative decline and absolute decline. CPFE: combined pulmonary fibrosis and emphysema; IPF: idiopathic pulmonary fibrosis; FVC: forced vital capacity. *=p<0.01 when comparing c). †=p<0.01 when comparing d).

Supplementary Table 7. DLco decline analysis in different subgroups of IPF patients

| Cohort | Subgroup | DLco data available cases/all case | Relative 1-year DLco decline (%) | | Absolute 1-year DLco decline (mls/year) |
|---|---|---|---|---|---|
| | | | Number of >15% (proportion) | Number of >10% (proportion) | Mean |
| Derivation cohort | Non-CPFE | 132/183 | 52 (39.39%) | 73 (55.30%) | 645.39 |
| | CPFE with emphysema <15% | 157/218 | 51 (32.48%) | 75 (47.77%) | 950.61 |
| | CPFE with emphysema ⩾15% | 75/99 | 33 (44.00%) | 44 (58.67%) | 954.13 |
| | Fibrosis-dominant CPFE | 140/191 | 48 (34.29%) | 67 (47.86%) | 957.04 |
| | Matched-CPFE | 92/126 | 36 (39.13%) | 52 (56.52%) | 943.68 |
| Replication cohort | Non-CPFE | 108/152 | 30 (27.78%) | 43 (39.81%) | 769.10 |
| | CPFE with emphysema <15% | 197/258 | 51 (25.89%) | 86 (43.65%) | 617.02 |
| | CPFE with emphysema ⩾15% | 81/100 | 29 (35.80%) | 45 (55.56%) | 561.34 |
| | Fibrosis-dominant CPFE | 175/227 | 48 (27.43%) | 81 (46.29%) | 623.83 |
| | Matched-CPFE | 103/131 | 32 (31.07%) | 50 (48.54%) | 561.68 |
| Combined drug trial cohort | Non-CPFE | 213/236 | 71 (33.33%) | 100 (46.95%) | 748.91 |
| | CPFE with emphysema <15% | 291/318 | 83 (28.52%) | 139 (47.77%) | 832.87 |
| | CPFE with emphysema ⩾15% | 93/100 | 37 (39.78%) | 53 (56.99%) | 883.39 |
| | Fibrosis-dominant CPFE | 260/281 | 79 (30.38%) | 128 (49.23%) | 844.65 |
| | Matched-CPFE | 124/137 | 41 (33.06%) | 64 (51.61%) | 846.06 |

The proportions of patients with more than 15% and 10% relative 1-year DLco decline, and the mean of absolute 1-year DLco decline in different subgroups in derivation and replication cohorts and the combined drug trial cohort (patients fulfilling criteria to enter IPF therapeutic trials in derivation and replication cohorts) are shown in this table. The number of subjects with available DLco decline versus the number of all subjects within a subgroup is shown in n/n format. We also compared a) non-CPFE with CPFE with emphysema ⩾15%, b) CPFE with emphysema ⩾15% and CPFE with emphysema <15%, c) non-CPFE with Matched-CPFE subtype, d) Fibrosis-dominant CPFE subtype and Matched-CPFE subtype in terms of the relative decline and absolute decline. No statistically significant between group differences were identified. CPFE: combined pulmonary fibrosis and emphysema; IPF: idiopathic pulmonary fibrosis; DLco: diffusion capacity for carbon monoxide.

Supplementary Table 8. Multivariable mixed-effects Cox proportional hazards regression models in non-CPFE patients and the two CPFE subgroups (10% emphysema threshold) who fulfill criteria to enter IPF therapeutic trials in combined derivation and replication IPF cohorts.

| Subgroup | Baseline severity and PFTs changes models | Entire study population | | | | | |
|---|---|---|---|---|---|---|---|
| | | N (N-observed) | C-index | P-value | Hazard ratio | 95% CI Lower | 95% CI Upper |
| Non-CPFE IPF patients | DLco % predicted<br>1-year FVC relative decline | 212 (87) | 0.812 | $2.63\times10^{-6}$<br>$1.29\times10^{-11}$ | 0.952<br>1.088 | 0.933<br>1.062 | 0.972<br>1.115 |
| | DLco % predicted<br>Binary 1-year FVC decline (5%) | 212 (87) | 0.805 | $9.97\times10^{-7}$<br>$9.94\times10^{-7}$ | 0.952<br>3.268 | 0.933<br>2.034 | 0.971<br>5.252 |
| | DLco % predicted<br>Binary 1-year FVC decline (10%) | 212 (87) | 0.807 | $1.40\times10^{-5}$<br>$2.13\times10^{-9}$ | 0.957<br>4.36 | 0.938<br>2.693 | 0.976<br>7.06 |
| | DLco % predicted<br>1-year DLco relative decline | 212 (87) | 0.800 | $7.88\times10^{-8}$<br>$4.25\times10^{-6}$ | 0.946<br>1.042 | 0.927<br>1.024 | 0.965<br>1.06 |
| | DLco % predicted<br>Binary 1-year DLco decline (10%) | 212 (87) | 0.805 | $5.09\times10^{-7}$<br>$6.23\times10^{-5}$ | 0.950<br>2.697 | 0.931<br>1.659 | 0.969<br>4.384 |
| | DLco % predicted<br>Binary 1-year DLco decline (15%) | 212 (87) | 0.808 | $4.65\times10^{-7}$<br>$5.74\times10^{-7}$ | 0.949<br>3.337 | 0.93<br>2.081 | 0.969<br>5.352 |
| CPFE patients with emphysema < 10% | DLco % predicted<br>1-year FVC relative decline | 233 (114) | 0.711 | $6.76\times10^{-8}$<br>$6.70\times10^{-7}$ | 0.954<br>1.049 | 0.938<br>1.03 | 0.971<br>1.069 |
| | DLco % predicted<br>Binary 1-year FVC decline (5%) | 233 (114) | 0.710 | $3.62\times10^{-8}$<br>0.0003 | 0.954<br>2.007 | 0.938<br>1.376 | 0.970<br>2.928 |
| | DLco % predicted<br>Binary 1-year FVC decline (10%) | 233 (114) | 0.699 | $2.53\times10^{-7}$<br>0.0001 | 0.955<br>2.282 | 0.939<br>1.502 | 0.972<br>3.469 |
| | DLco % predicted<br>1-year DLco relative decline | 233 (114) | 0.735 | $1.27\times10^{-9}$<br>$6.20\times10^{-9}$ | 0.948<br>1.04 | 0.931<br>1.027 | 0.964<br>1.054 |
| | DLco % predicted<br>Binary 1-year DLco decline (10%) | 233 (114) | 0.710 | $6.00\times10^{-9}$<br>0.0002 | 0.952<br>2.110 | 0.936<br>1.429 | 0.968<br>3.116 |
| | DLco % predicted<br>Binary 1-year DLco decline (15%) | 233 (114) | 0.719 | $6.84\times10^{-9}$<br>$5.87\times10^{-7}$ | 0.951<br>2.885 | 0.935<br>1.904 | 0.968<br>4.372 |
| CPFE patients with emphysema ⩾ 10% | DLco % predicted<br>1-year FVC relative decline | 144 (89) | 0.710 | $1.26\times10^{-6}$<br>0.0006 | 0.936<br>1.051 | 0.912<br>1.022 | 0.961<br>1.082 |
| | DLco % predicted<br>Binary 1-year FVC decline (5%) | 144 (89) | 0.700 | $3.23\times10^{-6}$<br>0.022 | 0.940<br>1.693 | 0.916<br>1.077 | 0.965<br>2.660 |
| | DLco % predicted<br>Binary 1-year FVC decline (10%) | 144 (89) | 0.708 | $4.02\times10^{-5}$<br>0.001 | 0.948<br>2.363 | 0.924<br>1.412 | 0.972<br>3.955 |
| | DLco % predicted<br>1-year DLco relative decline | 144 (89) | 0.723 | $6.70\times10^{-6}$<br>$5.45\times10^{-8}$ | 0.941<br>1.041 | 0.916<br>1.026 | 0.966<br>1.056 |
| | DLco % predicted<br>Binary 1-year DLco decline (10%) | 144 (89) | 0.691 | $3.07\times10^{-6}$<br>0.003 | 0.939<br>1.987 | 0.914<br>1.272 | 0.964<br>3.105 |
| | DLco % predicted<br>Binary 1-year DLco decline (15%) | 144 (89) | 0.730 | $2.43\times10^{-6}$<br>$2.33\times10^{-7}$ | 0.939<br>3.376 | 0.914<br>2.129 | 0.964<br>5.353 |

Multivariable mixed-effects Cox regression models were used to investigate associations with mortality for 1-year FVC decline and 1-year DLco decline after adjusting for patient age, gender, smoking status (never versus ever), antifibrotic use (never versus ever) and baseline disease severity estimated using DLco. Binary 1-year FVC decline uses 5% and 10% relative decline as thresholds, and binary 1-year DLco decline uses 10% and 15% relative decline as thresholds. Separate centres/countries within the derivation and replication cohorts were modelled as multilevel with random effects between centres/countries (a random intercept per centre/country). N: number of patients in mixed-effects Cox model. N-observed: number of deaths observed in N patients; C-index: concordance index; CPFE: combined pulmonary fibrosis and emphysema; IPF: idiopathic pulmonary fibrosis; FVC: forced vital capacity; DLco: diffusing capacity for carbon monoxide; CI: confidence interval.

Supplementary Table 9. Multivariable mixed-effects Cox proportional hazards regression models in non-CPFE patients and the two CPFE subgroups (15% emphysema threshold) in the derivation IPF cohorts.

| Subgroup | Baseline severity and PFTs changes models | Entire study population | | | | | |
|---|---|---|---|---|---|---|---|
| | | N (N-observed) | C-index | P-value | Hazard ratio | 95% CI | |
| | | | | | | Lower | Upper |
| Non-CPFE IPF patients | DLco % predicted<br>1-year FVC relative decline | 130 (61) | 0.821 | $1.94\times10^{-6}$<br>$3.02\times10^{-8}$ | 0.942<br>1.082 | 0.919<br>1.052 | 0.966<br>1.113 |
| | DLco % predicted<br>Binary 1-year FVC decline (5%) | 130 (61) | 0.805 | $7.65\times10^{-8}$<br>$1.09\times10^{-5}$ | 0.935<br>3.824 | 0.912<br>2.104 | 0.958<br>6.953 |
| | DLco % predicted<br>Binary 1-year FVC decline (10%) | 130 (61) | 0.811 | $4.33\times10^{-6}$<br>$4.96\times10^{-7}$ | 0.945<br>4.261 | 0.923<br>2.422 | 0.968<br>7.497 |
| | DLco % predicted<br>1-year DLco relative decline | 130 (61) | 0.803 | $1.62\times10^{-7}$<br>0.0001 | 0.937<br>1.038 | 0.914<br>1.018 | 0.960<br>1.058 |
| | DLco % predicted<br>Binary 1-year DLco decline (10%) | 130 (61) | 0.800 | $4.03\times10^{-7}$<br>0.0010 | 0.940<br>2.764 | 0.918<br>1.511 | 0.963<br>5.055 |
| | DLco % predicted<br>Binary 1-year DLco decline (15%) | 130 (61) | 0.811 | $9.11\times10^{-8}$<br>$4.69\times10^{-7}$ | 0.936<br>4.211 | 0.913<br>2.407 | 0.959<br>7.366 |
| CPFE patients with emphysema < 15% | DLco % predicted<br>1-year FVC relative decline | 149 (87) | 0.719 | $6.46\times10^{-9}$<br>0.0003 | 0.945<br>1.037 | 0.927<br>1.016 | 0.963<br>1.057 |
| | DLco % predicted<br>Binary 1-year FVC decline (5%) | 149 (87) | 0.722 | $2.52\times10^{-8}$<br>0.0002 | 0.949<br>2.487 | 0.931<br>1.546 | 0.966<br>4.001 |
| | DLco % predicted<br>Binary 1-year FVC decline (10%) | 149 (87) | 0.707 | $2.24\times10^{-7}$<br>0.016 | 0.953<br>1.847 | 0.935<br>1.122 | 0.970<br>3.039 |
| | DLco % predicted<br>1-year DLco relative decline | 149 (87) | 0.742 | $7.69\times10^{-10}$<br>$7.87\times10^{-6}$ | 0.939<br>1.038 | 0.920<br>1.021 | 0.958<br>1.055 |
| | DLco % predicted<br>Binary 1-year DLco decline (10%) | 149 (87) | 0.707 | $3.98\times10^{-8}$<br>0.0746 | 0.949<br>1.510 | 0.931<br>0.960 | 0.967<br>2.377 |
| | DLco % predicted<br>Binary 1-year DLco decline (15%) | 149 (87) | 0.725 | $4.72\times10^{-9}$<br>0.0009 | 0.946<br>2.213 | 0.929<br>1.380 | 0.964<br>3.548 |
| CPFE patients with emphysema ⩾ 15% | DLco % predicted<br>1-year FVC relative decline | 73 (49) | 0.729 | 0.0003<br>0.002 | 0.949<br>1.055 | 0.923<br>1.020 | 0.977<br>1.090 |
| | DLco % predicted<br>Binary 1-year FVC decline (5%) | 73 (49) | 0.723 | 0.0011<br>0.0202 | 0.957<br>2.169 | 0.932<br>1.128 | 0.983<br>4.170 |
| | DLco % predicted<br>Binary 1-year FVC decline (10%) | 73 (49) | 0.730 | 0.010<br>0.001 | 0.964<br>4.305 | 0.938<br>1.756 | 0.991<br>10.551 |
| | DLco % predicted<br>1-year DLco relative decline | 73 (49) | 0.742 | 0.0005<br>$7.28\times10^{-5}$ | 0.948<br>1.034 | 0.920<br>1.017 | 0.977<br>1.051 |
| | DLco % predicted<br>Binary 1-year DLco decline (10%) | 73 (49) | 0.720 | 0.0012<br>0.0566 | 0.956<br>1.842 | 0.930<br>0.983 | 0.982<br>3.451 |
| | DLco % predicted<br>Binary 1-year DLco decline (15%) | 73 (49) | 0.738 | 0.0008<br>0.0005 | 0.952<br>2.931 | 0.925<br>1.598 | 0.980<br>5.375 |

Multivariable mixed-effects Cox regression models were used to investigate associations with mortality for 1-year FVC decline and 1-year DLco decline after adjusting for patient age, gender, smoking status (never versus ever), antifibrotic use (never versus ever) and baseline disease severity estimated using DLco. Binary 1-year FVC decline uses 5% and 10% relative decline as thresholds, and binary 1-year DLco decline uses 10% and 15% relative decline as thresholds. Separate centres/countries within the derivation and replication cohorts were modelled as multilevel with random effects between centres/countries (a random intercept per centre/country). N: number of patients in mixed-effects Cox model. N-observed: number of deaths observed in N patients; C-index: concordance index; CPFE: combined pulmonary fibrosis and emphysema; IPF: idiopathic pulmonary fibrosis; FVC: forced vital capacity; DLco: diffusing capacity for carbon monoxide; CI: confidence interval.

Supplementary Table 10. Multivariable mixed-effects Cox proportional hazards regression models in non-CPFE patients and the two CPFE subgroups (15% emphysema threshold) in the replication IPF cohorts.

| Subgroup | Baseline severity and PFTs changes models | Entire study population | | | | | |
|---|---|---|---|---|---|---|---|
| | | N (N-observed) | C-index | P-value | Hazard ratio | 95% CI | |
| | | | | | | Lower | Upper |
| Non-CPFE IPF patients | DLco % predicted<br>1-year FVC relative decline | 108 (45) | 0.823 | $2.51\times10^{-5}$<br>$8.65\times10^{-5}$ | 0.940<br>1.086 | 0.913<br>1.042 | 0.967<br>1.132 |
| | DLco % predicted<br>Binary 1-year FVC decline (5%) | 108 (45) | 0.827 | $3.35\times10^{-5}$<br>0.002 | 0.942<br>2.719 | 0.916<br>1.425 | 0.969<br>5.187 |
| | DLco % predicted<br>Binary 1-year FVC decline (10%) | 108 (45) | 0.817 | $5.17\times10^{-5}$<br>0.004 | 0.944<br>2.733 | 0.918<br>1.374 | 0.971<br>5.437 |
| | DLco % predicted<br>1-year DLco relative decline | 108 (45) | 0.822 | $3.28\times10^{-6}$<br>0.019 | 0.933<br>1.032 | 0.906<br>1.005 | 0.960<br>1.059 |
| | DLco % predicted<br>Binary 1-year DLco decline (10%) | 108 (45) | 0.835 | $1.56\times10^{-5}$<br>0.013 | 0.938<br>2.373 | 0.911<br>1.201 | 0.966<br>4.688 |
| | DLco % predicted<br>Binary 1-year DLco decline (15%) | 108 (45) | 0.835 | $2.69\times10^{-5}$<br>0.006 | 0.941<br>2.693 | 0.915<br>1.336 | 0.968<br>5.428 |
| CPFE patients with emphysema < 15% | DLco % predicted<br>1-year FVC relative decline | 194 (102) | 0.750 | $1.93\times10^{-10}$<br>0.0005 | 0.944<br>1.053 | 0.927<br>1.023 | 0.961<br>1.085 |
| | DLco % predicted<br>Binary 1-year FVC decline (5%) | 194 (102) | 0.754 | $1.23\times10^{-10}$<br>0.0021 | 0.943<br>1.890 | 0.926<br>1.260 | 0.960<br>2.835 |
| | DLco % predicted<br>Binary 1-year FVC decline (10%) | 194 (102) | 0.760 | $1.85\times10^{-10}$<br>$2.44\times10^{-5}$ | 0.944<br>2.657 | 0.927<br>1.688 | 0.961<br>4.183 |
| | DLco % predicted<br>1-year DLco relative decline | 194 (102) | 0.776 | $3.01\times10^{-11}$<br>$4.21\times10^{-6}$ | 0.943<br>1.032 | 0.926<br>1.018 | 0.959<br>1.047 |
| | DLco % predicted<br>Binary 1-year DLco decline (10%) | 194 (102) | 0.766 | $8.95\times10^{-11}$<br>0.0002 | 0.944<br>2.181 | 0.928<br>1.454 | 0.961<br>3.272 |
| | DLco % predicted<br>Binary 1-year DLco decline (15%) | 194 (102) | 0.767 | $8.49\times10^{-10}$<br>$7.76\times10^{-6}$ | 0.946<br>2.798 | 0.929<br>1.782 | 0.963<br>4.393 |
| CPFE patients with emphysema ⩾ 15% | DLco % predicted<br>1-year FVC relative decline | 80 (51) | 0.722 | 0.001<br>0.122 | 0.952<br>1.027 | 0.923<br>0.993 | 0.981<br>1.063 |
| | DLco % predicted<br>Binary 1-year FVC decline (5%) | 80 (51) | 0.688 | 0.0031<br>0.8652 | 0.956<br>1.056 | 0.928<br>0.565 | 0.985<br>1.973 |
| | DLco % predicted<br>Binary 1-year FVC decline (10%) | 80 (51) | 0.706 | 0.007<br>0.079 | 0.959<br>2.052 | 0.930<br>0.920 | 0.988<br>4.576 |
| | DLco % predicted<br>1-year DLco relative decline | 80 (51) | 0.720 | 0.0003<br>0.01 | 0.946<br>1.026 | 0.917<br>1.006 | 0.975<br>1.047 |
| | DLco % predicted<br>Binary 1-year DLco decline (10%) | 80 (51) | 0.709 | 0.0002<br>0.0025 | 0.947<br>2.767 | 0.920<br>1.430 | 0.975<br>5.353 |
| | DLco % predicted<br>Binary 1-year DLco decline (15%) | 80 (51) | 0.724 | 0.0006<br>0.0003 | 0.950<br>3.846 | 0.922<br>1.866 | 0.978<br>7.925 |

Multivariable mixed-effects Cox regression models were used to investigate associations with mortality for 1-year FVC decline and 1-year DLco decline after adjusting for patient age, gender, smoking status (never versus ever), antifibrotic use (never versus ever) and baseline disease severity estimated using DLco. Binary 1-year FVC decline uses 5% and 10% relative decline as thresholds, and binary 1-year DLco decline uses 10% and 15% relative decline as thresholds. Separate centres/countries within the derivation and replication cohorts were modelled as multilevel with random effects between centres/countries (a random intercept per centre/country). N: number of patients in mixed-effects Cox model. N-observed: number of deaths observed in N patients; C-index: concordance index; CPFE: combined pulmonary fibrosis and emphysema; IPF: idiopathic pulmonary fibrosis; FVC: forced vital capacity; DLco: diffusing capacity for carbon monoxide; CI: confidence interval.

Supplementary Table 11. Multivariable mixed-effects Cox proportional hazards regression models in non-CPFE patients and the two CPFE subgroups (15% emphysema threshold) who fulfill criteria to enter IPF therapeutic trials in combined derivation and replication IPF cohorts.

| Subgroup | Baseline severity and PFTs changes models | Entire study population | | | | | |
|---|---|---|---|---|---|---|---|
| | | N (N-observed) | C-index | P-value | Hazard ratio | 95% CI | |
| | | | | | | Lower | Upper |
| Non-CPFE IPF patients | DLco % predicted<br>1-year FVC relative decline | 212 (87) | 0.812 | $2.63\times10^{-6}$<br>$1.29\times10^{-11}$ | 0.952<br>1.088 | 0.933<br>1.062 | 0.972<br>1.115 |
| | DLco % predicted<br>Binary 1-year FVC decline (5%) | 212 (87) | 0.805 | $9.97\times10^{-7}$<br>$9.94\times10^{-7}$ | 0.952<br>3.268 | 0.933<br>2.034 | 0.971<br>5.252 |
| | DLco % predicted<br>Binary 1-year FVC decline (10%) | 212 (87) | 0.807 | $1.40\times10^{-5}$<br>$2.13\times10^{-9}$ | 0.957<br>4.36 | 0.938<br>2.693 | 0.976<br>7.06 |
| | DLco % predicted<br>1-year DLco relative decline | 212 (87) | 0.800 | $7.88\times10^{-8}$<br>$4.25\times10^{-6}$ | 0.946<br>1.042 | 0.927<br>1.024 | 0.965<br>1.06 |
| | DLco % predicted<br>Binary 1-year DLco decline (10%) | 212 (87) | 0.805 | $5.09\times10^{-7}$<br>$6.23\times10^{-5}$ | 0.950<br>2.697 | 0.931<br>1.659 | 0.969<br>4.384 |
| | DLco % predicted<br>Binary 1-year DLco decline (15%) | 212 (87) | 0.808 | $4.65\times10^{-7}$<br>$5.74\times10^{-7}$ | 0.949<br>3.337 | 0.93<br>2.081 | 0.969<br>5.352 |
| CPFE patients with emphysema < 15% | DLco % predicted<br>1-year FVC relative decline | 285 (147) | 0.721 | $4.27\times10^{-11}$<br>$4.51\times10^{-7}$ | 0.948<br>1.045 | 0.933<br>1.028 | 0.963<br>1.064 |
| | DLco % predicted<br>Binary 1-year FVC decline (5%) | 285 (147) | 0.720 | $3.60\times10^{-11}$<br>0.0001 | 0.948<br>1.913 | 0.933<br>1.370 | 0.963<br>2.671 |
| | DLco % predicted<br>Binary 1-year FVC decline (10%) | 285 (147) | 0.714 | $2.96\times10^{-10}$<br>$6.63\times10^{-6}$ | 0.949<br>2.356 | 0.934<br>1.623 | 0.965<br>3.42 |
| | DLco % predicted<br>1-year DLco relative decline | 285 (147) | 0.760 | $1.71\times10^{-13}$<br>$5.28\times10^{-13}$ | 0.941<br>1.046 | 0.926<br>1.034 | 0.956<br>1.059 |
| | DLco % predicted<br>Binary 1-year DLco decline (10%) | 285 (147) | 0.730 | $4.37\times10^{-12}$<br>$1.50\times10^{-5}$ | 0.946<br>2.127 | 0.931<br>1.511 | 0.961<br>2.994 |
| | DLco % predicted<br>Binary 1-year DLco decline (15%) | 285 (147) | 0.739 | $1.74\times10^{-12}$<br>$2.99\times10^{-10}$ | 0.944<br>3.199 | 0.929<br>2.228 | 0.959<br>4.593 |
| CPFE patients with emphysema ⩾ 15% | DLco % predicted<br>1-year FVC relative decline | 92 (56) | 0.735 | 0.0001<br>0.0004 | 0.935<br>1.071 | 0.904<br>1.031 | 0.968<br>1.112 |
| | DLco % predicted<br>Binary 1-year FVC decline (5%) | 92 (56) | 0.722 | 0.0005<br>0.0253 | 0.944<br>2.030 | 0.913<br>1.091 | 0.975<br>3.777 |
| | DLco % predicted<br>Binary 1-year FVC decline (10%) | 92 (56) | 0.717 | 0.008<br>0.009 | 0.957<br>2.764 | 0.926<br>1.295 | 0.989<br>5.899 |
| | DLco % predicted<br>1-year DLco relative decline | 92 (56) | 0.714 | 0.0009<br>0.0009 | 0.945<br>1.029 | 0.914<br>1.012 | 0.977<br>1.047 |
| | DLco % predicted<br>Binary 1-year DLco decline (10%) | 92 (56) | 0.689 | 0.0009<br>0.0765 | 0.945<br>1.701 | 0.914<br>0.945 | 0.977<br>3.061 |
| | DLco % predicted<br>Binary 1-year DLco decline (15%) | 92 (56) | 0.720 | 0.002<br>0.001 | 0.948<br>2.623 | 0.917<br>1.478 | 0.98<br>4.657 |

Multivariable mixed-effects Cox regression models were used to investigate associations with mortality for 1-year FVC decline and 1-year DLco decline after adjusting for patient age, gender, smoking status (never versus ever), antifibrotic use (never versus ever) and baseline disease severity estimated using DLco. Binary 1-year FVC decline uses 5% and 10% relative decline as thresholds, and binary 1-year DLco decline uses 10% and 15% relative decline as thresholds. Separate centres/countries within the derivation and replication cohorts were modelled as multilevel with random effects between centres/countries (a random intercept per centre/country). N: number of patients in mixed-effects Cox model. N-observed: number of deaths observed in N patients; C-index: concordance index; CPFE: combined pulmonary fibrosis and emphysema; IPF: idiopathic pulmonary fibrosis; FVC: forced vital capacity; DLco: diffusing capacity for carbon monoxide; CI: confidence interval.

Supplementary Table 12. Multivariable mixed-effects Cox proportional hazards regression models in non-CPFE patients and the two CPFE SuStaIn subtypes in the derivation IPF cohorts.

| Subgroup | Baseline severity and PFTs changes models | Entire study population | | | | | |
|---|---|---|---|---|---|---|---|
| | | N (N-observed) | C-index | P-value | Hazard ratio | 95% CI | |
| | | | | | | Lower | Upper |
| Non-CPFE IPF patients | DLco % predicted<br>1-year FVC relative decline | 130 (61) | 0.821 | $1.94\times10^{-6}$<br>$3.02\times10^{-8}$ | 0.942<br>1.082 | 0.919<br>1.052 | 0.966<br>1.113 |
| | DLco % predicted<br>Binary 1-year FVC decline (5%) | 130 (61) | 0.805 | $7.65\times10^{-8}$<br>$1.09\times10^{-5}$ | 0.935<br>3.824 | 0.912<br>2.104 | 0.958<br>6.953 |
| | DLco % predicted<br>Binary 1-year FVC decline (10%) | 130 (61) | 0.811 | $4.33\times10^{-6}$<br>$4.96\times10^{-7}$ | 0.945<br>4.261 | 0.923<br>2.422 | 0.968<br>7.497 |
| | DLco % predicted<br>1-year DLco relative decline | 130 (61) | 0.803 | $1.62\times10^{-7}$<br>0.0001 | 0.937<br>1.038 | 0.914<br>1.018 | 0.960<br>1.058 |
| | DLco % predicted<br>Binary 1-year DLco decline (10%) | 130 (61) | 0.800 | $4.03\times10^{-7}$<br>0.0010 | 0.940<br>2.764 | 0.918<br>1.511 | 0.963<br>5.055 |
| | DLco % predicted<br>Binary 1-year DLco decline (15%) | 130 (61) | 0.811 | $9.11\times10^{-8}$<br>$4.69\times10^{-7}$ | 0.936<br>4.211 | 0.913<br>2.407 | 0.959<br>7.366 |
| Fibrosis-dominant CPFE patients | DLco % predicted<br>1-year FVC relative decline | 134 (76) | 0.731 | $1.31\times10^{-8}$<br>0.0005 | 0.943<br>1.039 | 0.924<br>1.017 | 0.962<br>1.062 |
| | DLco % predicted<br>Binary 1-year FVC decline (5%) | 134 (76) | 0.743 | $2.85\times10^{-8}$<br>$7.82\times10^{-5}$ | 0.947<br>2.765 | 0.928<br>1.669 | 0.965<br>4.580 |
| | DLco % predicted<br>Binary 1-year FVC decline (10%) | 134 (76) | 0.718 | $4.79\times10^{-7}$<br>0.009 | 0.952<br>2.018 | 0.934<br>1.189 | 0.970<br>3.424 |
| | DLco % predicted<br>1-year DLco relative decline | 134 (76) | 0.745 | $6.00\times10^{-9}$<br>0.0001 | 0.940<br>1.033 | 0.920<br>1.016 | 0.960<br>1.051 |
| | DLco % predicted<br>Binary 1-year DLco decline (10%) | 134 (76) | 0.719 | $1.08\times10^{-7}$<br>0.0831 | 0.948<br>1.540 | 0.929<br>0.945 | 0.967<br>2.509 |
| | DLco % predicted<br>Binary 1-year DLco decline (15%) | 134 (76) | 0.732 | $2.62\times10^{-8}$<br>0.003 | 0.946<br>2.168 | 0.928<br>1.313 | 0.965<br>3.577 |
| Matched-CPFE patients | DLco % predicted<br>1-year FVC relative decline | 88 (60) | 0.701 | 0.0003<br>0.0064 | 0.956<br>1.040 | 0.933<br>1.011 | 0.980<br>1.070 |
| | DLco % predicted<br>Binary 1-year FVC decline (5%) | 88 (60) | 0.704 | 0.0008<br>0.0589 | 0.960<br>1.711 | 0.938<br>0.980 | 0.983<br>2.987 |
| | DLco % predicted<br>Binary 1-year FVC decline (10%) | 88 (60) | 0.705 | 0.002<br>0.012 | 0.963<br>2.484 | 0.941<br>1.219 | 0.987<br>5.065 |
| | DLco % predicted<br>1-year DLco relative decline | 88 (60) | 0.727 | 0.0006<br>$1.07\times10^{-5}$ | 0.957<br>1.036 | 0.933<br>1.020 | 0.981<br>1.053 |
| | DLco % predicted<br>Binary 1-year DLco decline (10%) | 88 (60) | 0.688 | 0.0011<br>0.0699 | 0.961<br>1.674 | 0.939<br>0.959 | 0.984<br>2.922 |
| | DLco % predicted<br>Binary 1-year DLco decline (15%) | 88 (60) | 0.721 | 0.001<br>0.0004 | 0.961<br>2.634 | 0.938<br>1.535 | 0.984<br>4.518 |

Multivariable mixed-effects Cox regression models were used to investigate associations with mortality for 1-year FVC decline and 1-year DLco decline after adjusting for patient age, gender, smoking status (never versus ever), antifibrotic use (never versus ever) and baseline disease severity estimated using DLco. Binary 1-year FVC decline uses 5% and 10% relative decline as thresholds, and binary 1-year DLco decline uses 10% and 15% relative decline as thresholds. Separate centres/countries within the derivation and replication cohorts were modelled as multilevel with random effects between centres/countries (a random intercept per centre/country). N: number of patients in mixed-effects Cox model. N-observed: number of deaths observed in N patients; C-index: concordance index; CPFE: combined pulmonary fibrosis and emphysema; IPF: idiopathic pulmonary fibrosis; FVC: forced vital capacity; DLco: diffusing capacity for carbon monoxide; CI: confidence interval.

Supplementary Table 13. Multivariable mixed-effects Cox proportional hazards regression models in non-CPFE patients and the two CPFE SuStaIn subtypes in the replication IPF cohorts.

| Subgroup | Baseline severity and PFTs changes models | Entire study population | | | | | |
|---|---|---|---|---|---|---|---|
| | | N (N-observed) | C-index | P-value | Hazard ratio | 95% CI Lower | 95% CI Upper |
| Non-CPFE IPF patients | DLco % predicted<br>1-year FVC relative decline | 108 (45) | 0.823 | $2.51\times10^{-5}$<br>$8.65\times10^{-5}$ | 0.940<br>1.086 | 0.913<br>1.042 | 0.967<br>1.132 |
| | DLco % predicted<br>Binary 1-year FVC decline (5%) | 108 (45) | 0.827 | $3.35\times10^{-5}$<br>0.002 | 0.942<br>2.719 | 0.916<br>1.425 | 0.969<br>5.187 |
| | DLco % predicted<br>Binary 1-year FVC decline (10%) | 108 (45) | 0.817 | $5.17\times10^{-5}$<br>0.004 | 0.944<br>2.733 | 0.918<br>1.374 | 0.971<br>5.437 |
| | DLco % predicted<br>1-year DLco relative decline | 108 (45) | 0.822 | $3.28\times10^{-6}$<br>0.019 | 0.933<br>1.032 | 0.906<br>1.005 | 0.960<br>1.059 |
| | DLco % predicted<br>Binary 1-year DLco decline (10%) | 108 (45) | 0.835 | $1.56\times10^{-5}$<br>0.013 | 0.938<br>2.373 | 0.911<br>1.201 | 0.966<br>4.688 |
| | DLco % predicted<br>Binary 1-year DLco decline (15%) | 108 (45) | 0.835 | $2.69\times10^{-5}$<br>0.006 | 0.941<br>2.693 | 0.915<br>1.336 | 0.968<br>5.428 |
| Fibrosis-dominant CPFE patients | DLco % predicted<br>1-year FVC relative decline | 173 (95) | 0.764 | $2.26\times10^{-11}$<br>0.0008 | 0.938<br>1.051 | 0.921<br>1.021 | 0.956<br>1.082 |
| | DLco % predicted<br>Binary 1-year FVC decline (5%) | 173 (95) | 0.765 | $2.71\times10^{-11}$<br>0.0095 | 0.939<br>1.750 | 0.921<br>1.147 | 0.956<br>2.671 |
| | DLco % predicted<br>Binary 1-year FVC decline (10%) | 173 (95) | 0.770 | $2.11\times10^{-11}$<br>0.0003 | 0.938<br>2.396 | 0.921<br>1.497 | 0.956<br>3.836 |
| | DLco % predicted<br>1-year DLco relative decline | 173 (95) | 0.782 | $8.97\times10^{-12}$<br>$9.06\times10^{-5}$ | 0.939<br>1.028 | 0.922<br>1.014 | 0.956<br>1.042 |
| | DLco % predicted<br>Binary 1-year DLco decline (10%) | 173 (95) | 0.772 | 0.0029<br>$1.82\times10^{-10}$ | 1.890<br>0.941 | 1.244<br>0.924 | 2.873<br>0.959 |
| | DLco % predicted<br>Binary 1-year DLco decline (15%) | 173 (95) | 0.772 | $1.82\times10^{-10}$<br>0.0003 | 0.941<br>2.363 | 0.924<br>1.480 | 0.959<br>3.771 |
| Matched-CPFE patients | DLco % predicted<br>1-year FVC relative decline | 101 (58) | 0.719 | $3.61\times10^{-5}$<br>0.226 | 0.942<br>1.021 | 0.915<br>0.987 | 0.969<br>1.056 |
| | DLco % predicted<br>Binary 1-year FVC decline (5%) | 101 (58) | 0.708 | $7.26\times10^{-5}$<br>0.7189 | 0.945<br>1.112 | 0.919<br>0.624 | 0.972<br>1.982 |
| | DLco % predicted<br>Binary 1-year FVC decline (10%) | 101 (58) | 0.729 | 0.0001<br>0.021 | 0.947<br>2.361 | 0.921<br>1.137 | 0.975<br>4.906 |
| | DLco % predicted<br>1-year DLco relative decline | 101 (58) | 0.745 | $7.93\times10^{-6}$<br>0.0013 | 0.937<br>1.033 | 0.911<br>1.013 | 0.964<br>1.054 |
| | DLco % predicted<br>Binary 1-year DLco decline (10%) | 101 (58) | 0.747 | $8.24\times10^{-6}$<br>0.0001 | 0.941<br>3.468 | 0.916<br>1.845 | 0.967<br>6.517 |
| | DLco % predicted<br>Binary 1-year DLco decline (15%) | 101 (58) | 0.764 | $2.09\times10^{-5}$<br>$1.33\times10^{-5}$ | 0.943<br>4.858 | 0.917<br>2.385 | 0.969<br>9.895 |

Multivariable mixed-effects Cox regression models were used to investigate associations with mortality for 1-year FVC decline and 1-year DLco decline after adjusting for patient age, gender, smoking status (never versus ever), antifibrotic use (never versus ever) and baseline disease severity estimated using DLco. Binary 1-year FVC decline uses 5% and 10% relative decline as thresholds, and binary 1-year DLco decline uses 10% and 15% relative decline as thresholds. Separate centres/countries within the derivation and replication cohorts were modelled as multilevel with random effects between centres/countries (a random intercept per centre/country). N: number of patients in mixed-effects Cox model. N-observed: number of deaths observed in N patients; C-index: concordance index; CPFE: combined pulmonary fibrosis and emphysema; IPF: idiopathic pulmonary fibrosis; FVC: forced vital capacity; DLco: diffusing capacity for carbon monoxide; CI: confidence interval.

Supplementary Table 14. Multivariable mixed-effects Cox proportional hazards regression models in non-CPFE patients and the two CPFE SuStaIn subtypes who fulfill criteria to enter IPF therapeutic trials in combined derivation and replication IPF cohorts.

| Subgroup | Baseline severity and PFTs changes models | Entire study population | | | | | |
|---|---|---|---|---|---|---|---|
| | | N (N-observed) | C-index | P-value | Hazard ratio | 95% CI | |
| | | | | | | Lower | Upper |
| Non-CPFE IPF patients | DLco % predicted | 212 (87) | 0.812 | $2.63\times10^{-6}$ | 0.952 | 0.933 | 0.972 |
| | 1-year FVC relative decline | | | $1.29\times10^{-11}$ | 1.088 | 1.062 | 1.115 |
| | DLco % predicted | 212 (87) | 0.805 | $9.97\times10^{-7}$ | 0.952 | 0.933 | 0.971 |
| | Binary 1-year FVC decline (5%) | | | $9.94\times10^{-7}$ | 3.268 | 2.034 | 5.252 |
| | DLco % predicted | 212 (87) | 0.807 | $1.40\times10^{-5}$ | 0.957 | 0.938 | 0.976 |
| | Binary 1-year FVC decline (10%) | | | $2.13\times10^{-9}$ | 4.36 | 2.693 | 7.06 |
| | DLco % predicted | 212 (87) | 0.800 | $7.88\times10^{-8}$ | 0.946 | 0.927 | 0.965 |
| | 1-year DLco relative decline | | | $4.25\times10^{-6}$ | 1.042 | 1.024 | 1.06 |
| | DLco % predicted | 212 (87) | 0.805 | $5.09\times10^{-7}$ | 0.950 | 0.931 | 0.969 |
| | Binary 1-year DLco decline (10%) | | | $6.23\times10^{-5}$ | 2.697 | 1.659 | 4.384 |
| | DLco % predicted | 212 (87) | 0.808 | $4.65\times10^{-7}$ | 0.949 | 0.93 | 0.969 |
| | Binary 1-year DLco decline (15%) | | | $5.74\times10^{-7}$ | 3.337 | 2.081 | 5.352 |
| Fibrosis-dominant CPFE patients | DLco % predicted | 255 (131) | 0.727 | $9.64\times10^{-11}$ | 0.947 | 0.932 | 0.963 |
| | 1-year FVC relative decline | | | $5.19\times10^{-6}$ | 1.045 | 1.025 | 1.064 |
| | DLco % predicted | 255 (131) | 0.730 | $8.67\times10^{-11}$ | 0.948 | 0.932 | 0.963 |
| | Binary 1-year FVC decline (5%) | | | 0.0005 | 1.877 | 1.319 | 2.671 |
| | DLco % predicted | 255 (131) | 0.721 | $5.41\times10^{-10}$ | 0.949 | 0.933 | 0.965 |
| | Binary 1-year FVC decline (10%) | | | $6.12\times10^{-5}$ | 2.243 | 1.511 | 3.331 |
| | DLco % predicted | 255 (131) | 0.759 | $1.94\times10^{-12}$ | 0.942 | 0.926 | 0.958 |
| | 1-year DLco relative decline | | | $3.37\times10^{-10}$ | 1.042 | 1.028 | 1.055 |
| | DLco % predicted | 255 (131) | 0.734 | $1.28\times10^{-11}$ | 0.945 | 0.930 | 0.961 |
| | Binary 1-year DLco decline (10%) | | | 0.0001 | 2.028 | 1.417 | 2.901 |
| | DLco % predicted | 255 (131) | 0.741 | $1.19\times10^{-11}$ | 0.945 | 0.930 | 0.961 |
| | Binary 1-year DLco decline (15%) | | | $9.46\times10^{-9}$ | 3.009 | 2.066 | 4.384 |
| Matched-CPFE patients | DLco % predicted | 122 (72) | 0.696 | $8.17\times10^{-5}$ | 0.943 | 0.916 | 0.971 |
| | 1-year FVC relative decline | | | 0.0006 | 1.058 | 1.025 | 1.093 |
| | DLco % predicted | 122 (72) | 0.680 | 0.0002 | 0.947 | 0.921 | 0.974 |
| | Binary 1-year FVC decline (5%) | | | 0.0509 | 1.663 | 0.998 | 2.772 |
| | DLco % predicted | 122 (72) | 0.686 | 0.002 | 0.957 | 0.930 | 0.984 |
| | Binary 1-year FVC decline (10%) | | | 0.002 | 2.669 | 1.420 | 5.015 |
| | DLco % predicted | 122 (72) | 0.722 | 0.0001 | 0.944 | 0.917 | 0.972 |
| | 1-year DLco relative decline | | | $1.39\times10^{-7}$ | 1.041 | 1.025 | 1.056 |
| | DLco % predicted | 122 (72) | 0.684 | $8.40\times10^{-5}$ | 0.944 | 0.917 | 0.971 |
| | Binary 1-year DLco decline (10%) | | | 0.0007 | 2.412 | 1.453 | 4.006 |
| | DLco % predicted | 122 (72) | 0.730 | 0.0004 | 0.948 | 0.921 | 0.977 |
| | Binary 1-year DLco decline (15%) | | | $9.58\times10^{-7}$ | 3.606 | 2.159 | 6.023 |

Multivariable mixed-effects Cox regression models were used to investigate associations with mortality for 1-year FVC decline and 1-year DLco decline after adjusting for patient age, gender, smoking status (never versus ever), antifibrotic use (never versus ever) and baseline disease severity estimated using DLco. Binary 1-year FVC decline uses 5% and 10% relative decline as thresholds, and binary 1-year DLco decline uses 10% and 15% relative decline as thresholds. Separate centres/countries within the derivation and replication cohorts were modelled as multilevel with random effects between centres/countries (a random intercept per centre/country). N: number of patients in mixed-effects Cox model. N-observed: number of deaths observed in N patients; C-index: concordance index; CPFE: combined pulmonary fibrosis and emphysema; IPF: idiopathic pulmonary fibrosis; FVC: forced vital capacity; DLco: diffusing capacity for carbon monoxide; CI: confidence interval.

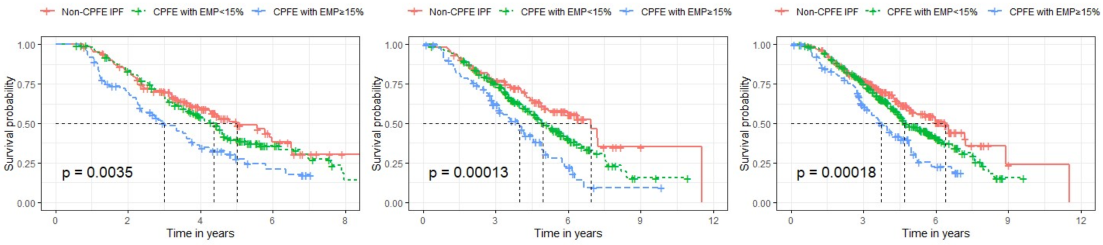


(a) (b) (c)

Supplementary Figure 1. Kaplan-Meier curves of non-CPFE IPF patients (red), CPFE patients with emphysema <15% (green) and CPFE patients with emphysema ⩾ 15% (blue) in the derivation cohort (a), the replication cohort (b), combined derivation and replication cohort patients qualifying for therapeutic trials (c). Log-rank tests show a significant difference in mortality between the three subtypes in all three analyses.

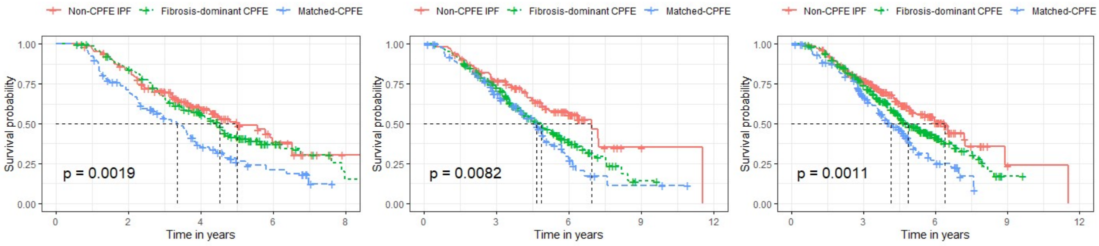


(a) (b) (c)

Supplementary Figure 2. Kaplan-Meier curves of non-CPFE IPF patients (red), fibrosis-dominant CPFE patients (green) and Matched-CPFE patients (blue) in the derivation cohort (a), the replication cohort (b), combined derivation and replication cohort patients qualifying for therapeutic trials (c). Log-rank tests show a significant difference in mortality between the three subtypes in all three analyses.

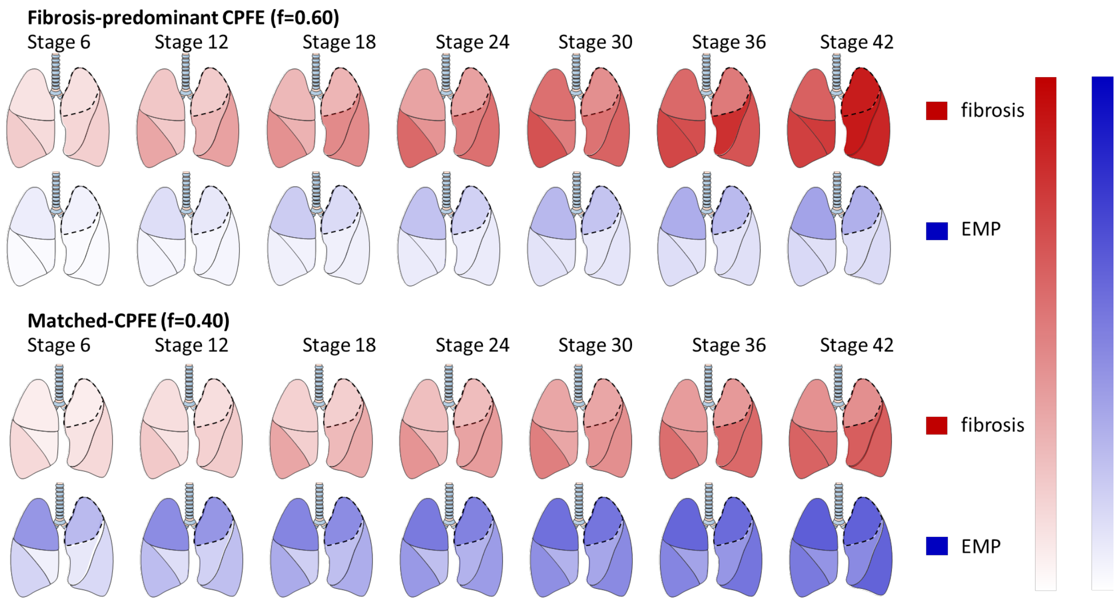


Supplementary Figure 3. Identification of CPFE subtypes and subtype disease progression modelled by SuStaIn in the derivation cohort. The rows show progression patterns of fibrosis extent (in red) and emphysema extent (in blue) in 6 lung zones (upper, middle and lower) in the two CPFE subtypes identified by SuStaIn: fibrosis-dominant CPFE and Matched-CPFE. Seven disease stages are highlighted, expressed as z-score intervals, In the fibrosis-dominant CPFE subtype comprising 60% of the cohort (top two rows), fibrosis is more severe at an early stage followed by a later emergence of emphysema. In the Matched-CPFE subtype comprising 40% of the cohort (bottom two rows), fibrosis and emphysema get worse together, with later stages showing relatively more extensive emphysema and less fibrosis compared to the fibrosis-dominant CPFE subtype. The upper lobe predominance of emphysema seen at early disease stages no longer exists in the later stages of the Matched-CPFE subtype. CPFE: Combined pulmonary fibrosis and emphysema. This figure was produced with the assistance of Servier Medical Art (https://smart.servier.com).

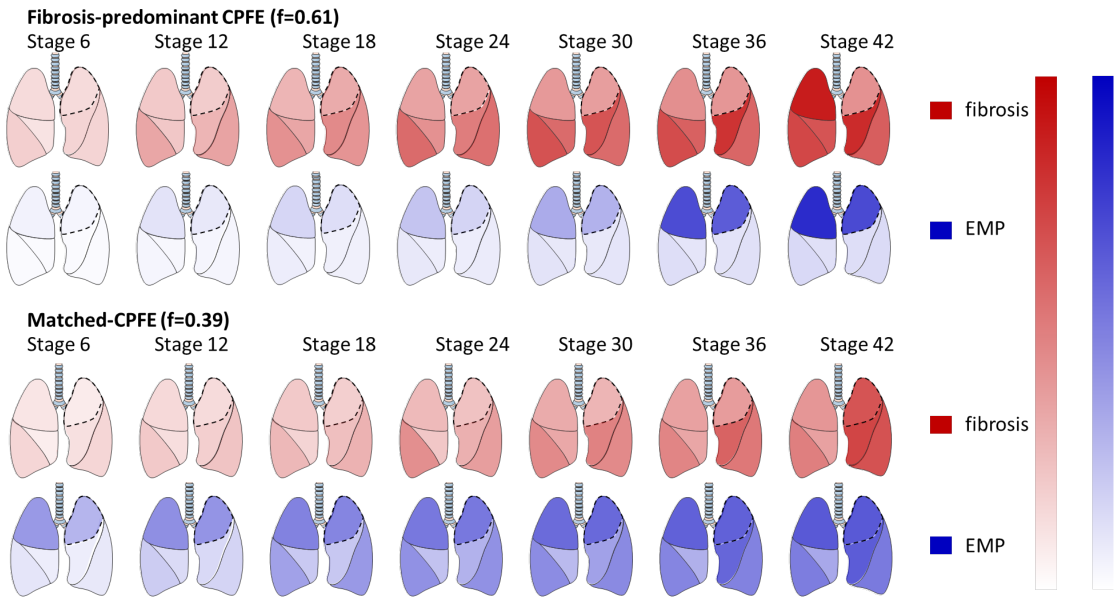


Supplementary Figure 4. Identification of CPFE subtypes and subtype disease progression modelled by SuStaIn in the replication cohort. The rows show progression patterns of fibrosis extent (in red) and emphysema extent (in blue) in 6 lung zones (upper, middle and lower) in the two CPFE subtypes identified by SuStaIn: Fibrosis-dominant CPFE and Matched-CPFE. Seven disease stages are highlighted, expressed as z-score intervals, In the fibrosis-dominant CPFE subtype comprising 61% of the cohort (top two rows), fibrosis is more severe at an early stage followed by a later emergence of emphysema. In the Matched-CPFE subtype comprising 39% of the cohort (bottom two rows), fibrosis and emphysema get worse together, with later stages showing relatively more extensive emphysema and less fibrosis compared to the fibrosis-dominant CPFE subtype. The upper lobe predominance of emphysema seen at early disease stages no longer exists in the later stages of the Matched-CPFE subtype. CPFE: Combined pulmonary fibrosis and emphysema. This figure was produced with the assistance of Servier Medical Art (https://smart.servier.com).
